# Increasing SARS-CoV-2 antibody prevalence in England at the start of the second wave: REACT-2 Round 4 cross-sectional study in 160,000 adults

**DOI:** 10.1101/2021.07.21.21260926

**Authors:** Helen Ward, Christina Atchison, Matt Whitaker, Christl A Donnelly, Steven Riley, Deborah Ashby, Ara Darzi, Wendy S Barclay, Graham Cooke, Paul Elliott, for the REACT study team

## Abstract

**Background:** REACT-2 Study 5 is a population survey of the prevalence of SARS-CoV-2 antibodies in the community in England.

**Methods:** We contacted a random sample of the population by sending a letter to named individuals aged 18 or over from the NHS GP registrations list. We then sent respondents a lateral flow immunoassay (LFIA) kit for SARS-CoV-2 antibody self-testing and asked them to perform the test at home and complete a questionnaire, including reporting of their test result. Overall, 161,537 adults completed questionnaires and self-administered LFIA tests for IgG against SARS-CoV-2 between 27 October and 10 November 2020.

**Results:** The overall adjusted and weighted prevalence was 5.6% (95% CI 5.4-5.7). This was an increase from 4.4% (4.3-4.5) in round 3 (September), a relative increase of 26.9% (24.0-29.9).The largest increase by age was in the 18 to 24 year old age group, which increased (adjusted and weighted) from 6.7% (6.3-7.2) to 9.9% (9.3-10.4), and in students, (adjusted, unweighted) from 5.9% (4.8-7.1) to 12.1% (10.8-13.5). Prevalence increased most in Yorkshire and The Humber, from 3.4% (3.0-3.8) to 6.3% (5.9-6.8) and the North West from 4.5% (4.2-4.9) to 7.7% (7.2-8.1). In contrast, the prevalence in London was stable, at 9.5% (9.0-9.9) and 9.5% (9.1-10.0) in rounds 3 and 4 respectively. We found the highest prevalence in people of Bangladeshi 15.1% (10.9-20.5), Pakistani 13.9% (11.2-17.2) and African 13.5% (10.7-16.8) ethnicity, and lowest in those of white British ethnicity at 4.2% (4.0-4.3).

**Interpretation:** The second wave of infection in England is apparent in increasing antibody prevalence, particularly in younger people, students, and in the Northern Regions. By late October a large proportion of the population remained susceptible to SARS-CoV-2 infection in England based on naturally acquired immunity from the first and early second wave.

## Introduction

Repeated prevalence surveys of SARS-CoV-2 antibodies can provide information on the distribution of infection within the population.[1,2] Antibody prevalence will change over time dependent on the incidence of infection, uptake of vaccination and waning of detectable antibody levels in those previously infected. England had a large first wave of COVID-19 in March and April 2020, leading to high mortality,[3] and significant morbidity for those with long COVID.[4] There was a national lockdown with the closure of schools, universities, hospitality, all but essential retail, and advice to work from home and avoid non-essential travel from late March with relaxations starting in May which led to a sustained reduction in cases and deaths until the second wave started in late August.[5]

We previously reported antibody positivity based on a self-administered lateral flow immunoassay (LFIA) test from a representative survey of 105,000 individuals during June and July 2020 where we found an overall prevalence, weighted to the adult population of England, of 6.0%.[6] Over two subsequent rounds of study in August and September 2020 we observed a decline in positivity to 4.4% which we attribute to waning antibody levels.[7] Here we report the prevalence of detectable antibody in a further round in October to November 2020 as part of the REal-time assessment of Community Transmission-2 (REACT-2) programme.[8]

## Methods

The REACT study protocol has been published.[8] Briefly, we included non-overlapping community samples from the adult population 18 years and older, using a self-administered questionnaire and LFIA test at home (Table 1).[6,8] Invitations were sent to named individuals randomly selected from the National Health Service (NHS) patient list which includes anyone registered with a General Practitioner (primary care physician) in England, covering almost the entire population. We aimed for a sample size of 150,000 in round 4 to obtain prevalence estimates for the 315 local authorities in England. Registration was closed after 200,721 people signed up, which was 36% of those invited registered; 169,927 completed a survey and 161,537 (29% of those invited, 80% of those registered) provided a valid (IgG positive or negative) antibody result (Supplementary Table S1). Those who registered were posted a self-administered point-of-care LFIA test (Fortress Diagnostics, Northern Ireland) with written and video instructions. The assay uses the S1 subunit (including RBD). The sensitivity of finger-prick blood (self-read) for IgG antibodies was 84.4% (70.5, 93.5) in RT-PCR confirmed cases in healthcare workers, and specificity 98.6% (97.1, 99.4) in pre-pandemic sera.[9] Participants completed a short registration questionnaire (online/telephone) and a further survey upon completion of their self-test. Survey instruments are available on the study website: (https://www.imperial.ac.uk/medicine/research-and-impact/groups/react-study/).

**Table 1:** REACT-2 rounds 1-4: community prevalence of IgG antibodies to SARS-CoV-2 in adults in England, adjusted and weighted, June 2020 – Nov 2020

|  | Total antibody positive | Total tests (with valid results) | Crude prevalence [95% confidence intervals] | Adjusted and weighted<sup>1</sup> prevalence [95% confidence intervals] |
| --- | --- | --- | --- | --- |
| Round 1 (20 Jun - 13 July) | 5544 | 99908 | 5.55 [5.41-5.69] | 5.96 [5.78-6.14] |
| Round 2 (31 Jul - 13 Aug) | 4995 | 105829 | 4.72 [4.59-4.85] | 4.83 [4.67-5.00] |
| Round 3 (15 Sept - 28 Sept) | 7037 | 159367 | 4.42 [4.32-4.52] | 4.38 [4.25-4.51] |
| Round 4 (27 Oct - 10 Nov) | 8431 | 161537 | 5.22 [5.11-5.33] | 5.56 [5.43-5.71] |

<sup>1</sup> Adjusted for test performance, weighted by age, sex, region, deprivation, ethnicity

The prevalence from each round was calculated as the proportion of individuals reporting a valid test result with a positive IgG result, adjusted for test performance,[10] and weighted at national level for age, sex, region, ethnicity and deprivation to the adult population of England.[6] Index of Multiple Deprivation 2019 (IMD) was used as a measure of relative deprivation, based on seven domains at a small local area level across England (income, employment, education, health, crime, barriers to housing and services, and living environment).[11] Epidemic curves were constructed retrospectively using information from participants with a positive antibody test who reported date of onset for a confirmed or possible case of COVID-19, i.e. excluding those who were asymptomatic and for whom date of onset was unknown. Logistic regression models were developed for prevalence by key covariates, and adjusted for age, sex and region, and additionally for ethnicity, deprivation, household size and occupation. We used complete case analysis without imputation. Confidence intervals for the changes in prevalence were calculated using a normal approximation to the sampling distribution of a difference in prevalences.[12]

Data were analysed using the statistical package R version 4.0.0.[13]

We obtained research ethics approval from the South Central-Berkshire B Research Ethics Committee (IRAS ID: 283787), and Medicines and Healthcare products Regulatory Agency approval for use of the LFIA for research purposes only. A REACT Public Advisory Group provides input into the design and conduct of the research.

## Results

Results were available from 161,537 people tested between 27 October and 10 November 2020. Over half of these test results were submitted in the first three days, i.e. by 29 October. The overall adjusted and weighted prevalence was 5.6% (95% CI 5.4-5.7). This was an increase from 4.4% (4.3-4.5) in round 3 (September), a relative increase of 26.9% (24.0-29.9). The results over the first 4 rounds show the highest prevalence in the first round in June, declining over the next 2 rounds and then increasing again by the end of October (Table 1).

Figure 1(a) is a reconstructed epidemic curve, based on date of onset reported by those with positive antibody tests who reported a confirmed or suspected case of COVID-19, with each round shown. It clearly shows that, from all rounds, most of the cases were from the first wave in March and April 2020; it also shows the start of the second wave becoming apparent in early September in the plot from round 4. The shaded area is where there will be under-ascertainment of cases as this IgG antibody test is validated for detection 21 or more days post onset.[9] Figure 1(b) shows the same reconstruction by age group, and demonstrates a marked increase from early September in those aged 18 to 24, with slower and slightly later increases in older age groups.

**Figure 1:**
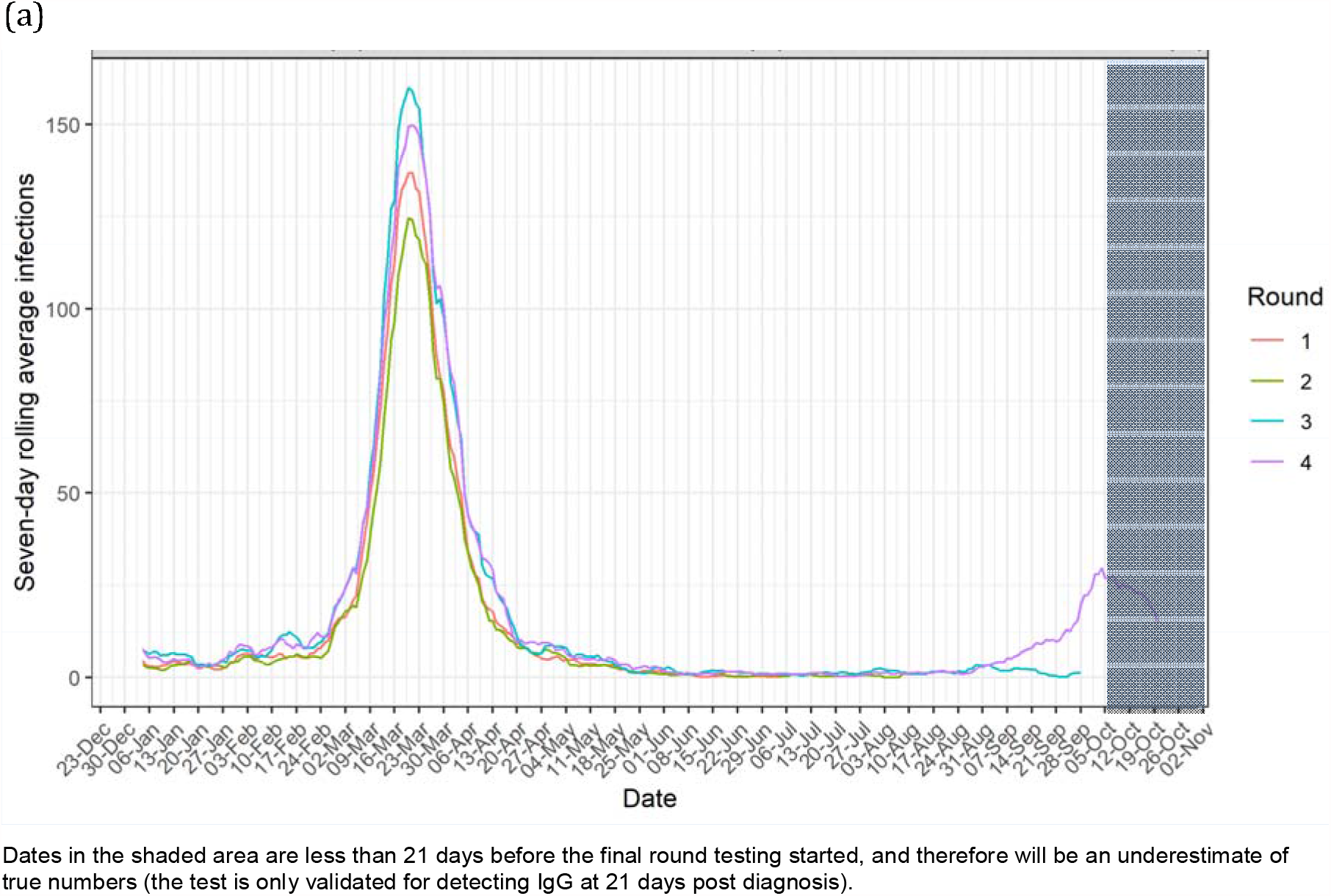
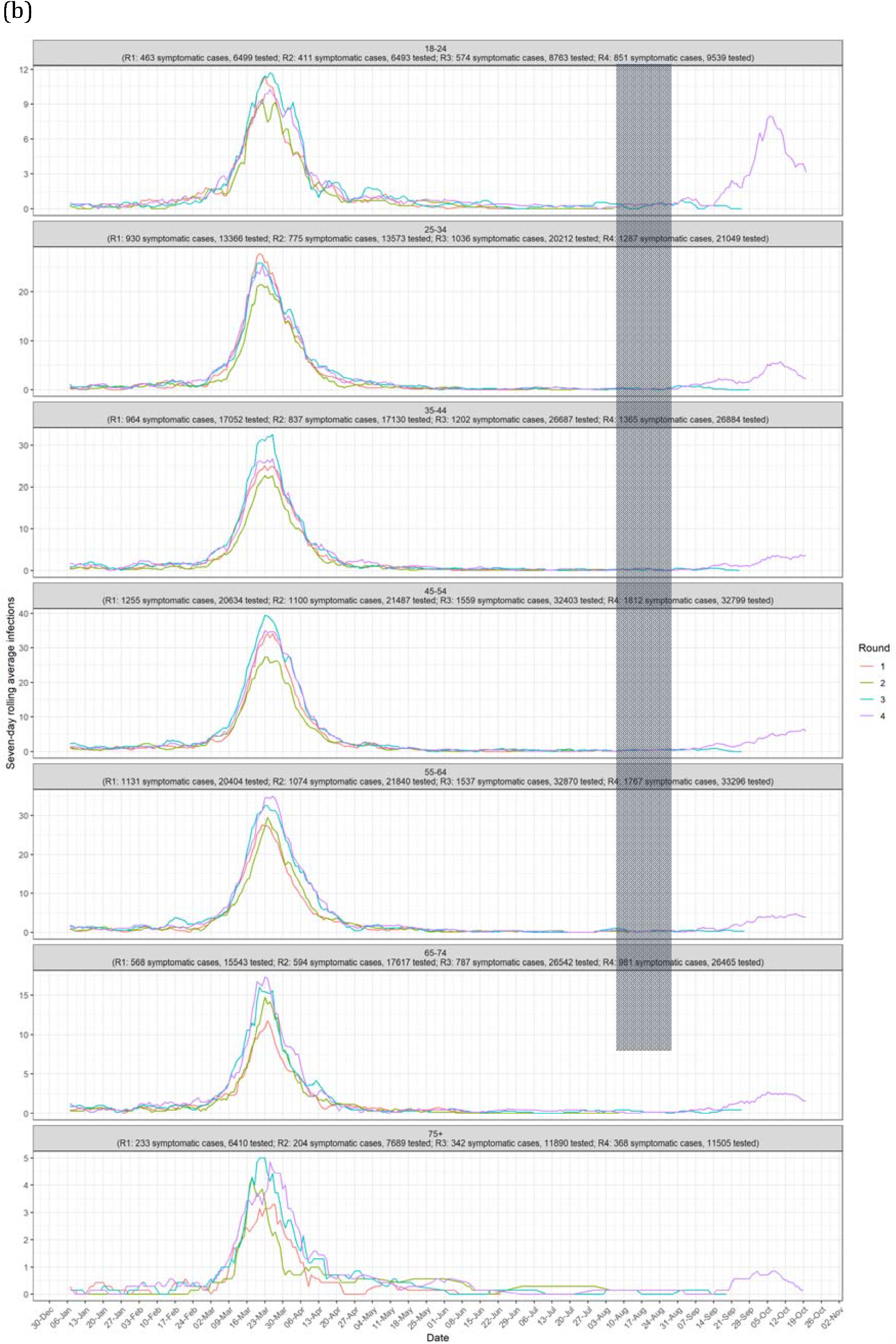
REACT-2 rounds 1-4: reconstructed epidemic curve from number of symptomatic infections per week, by date of onset in antibody positive participants reporting symptoms (a) all, (b) by age group

The characteristics of the early second wave are suggested by the relative changes in prevalence between rounds 3 and 4 in different groups. The largest increase by age was in the 18 to 24 year old age group, which increased (adjusted and weighted) from 6.7% (6.3-7.2) to 9.9% (9.3-10.4), in students, (adjusted, unweighted) from 5.9% (4.8-7.1) to 12.1% (10.8-13.5). Prevalence increased most in Yorkshire and The Humber, from 3.4% (3.0-3.8) to 6.3% (5.9-6.8) and the North West from 4.5% (4.2-4.9) to 7.7% (7.2-8.1), with higher rates in some lower tier local authority (LTLA) areas, including Liverpool and Knowsley, both of which are in the top five LTLAs ranked by prevalence (Supplementary Table S5). In contrast, the prevalence in London was stable, at 9.5% (9.0-9.9) and 9.5% (9.1-10.0) in rounds 3 and 4 respectively. There was little or no change in prevalence in people of Black (African, Caribbean, British and other) ethnicity, and in health and care home workers.

Figure 2 shows results of logistic regression for round 4, with multivariable adjusted odds ratios shown in a forest plot. It shows similar associations to earlier rounds,[6,7] but with additional covariates of interest, including increased odds of antibody positivity for students. Figure 2 also shows the association of antibody prevalence with household income, with a trend in the opposite direction to that for area level deprivation. It also shows households with children have lower odds of antibody positivity than those without. Supplementary Table S2 shows a breakdown of prevalence by ethnic sub-categories, showing the highest prevalence in people of Bangladeshi 15.1% (10.9-20.5), Pakistani13.9% (11.2-17.2) and African 13.5% (10.7-16.8) ethnicity, and lowest in those of White British ethnicity at 4.2% (4.0-4.3).

**Figure 2:**
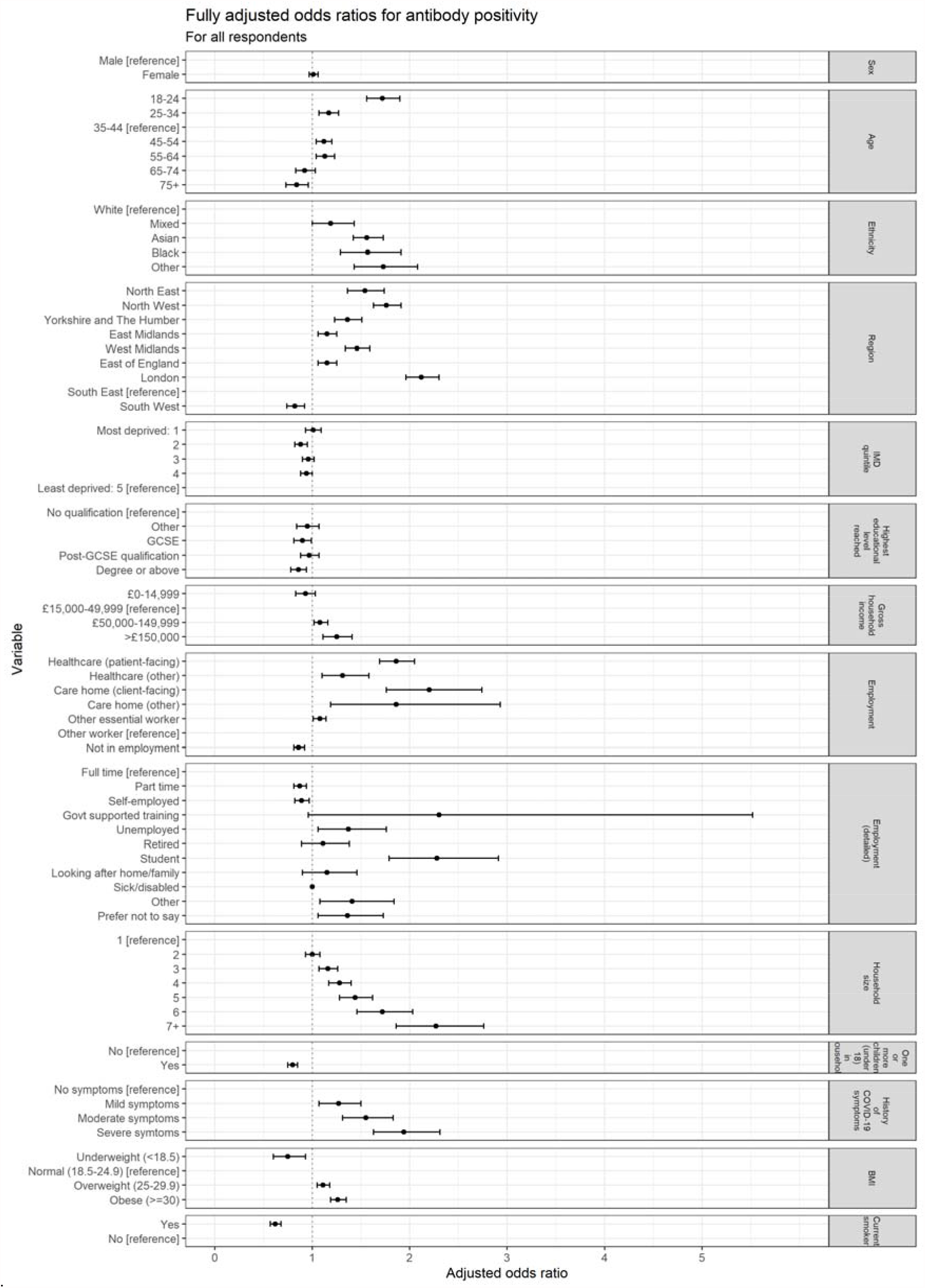
REACT-2 round 4: logistic regression for SARS-CoV-2 antibodies: adjusted Odds Ratios (95% CI) for sociodemographic and clinical covariates **Legend:** Jointly adjusted odds ratios [95% confidence intervals] from multivariable logistic regression for the covariate of interest. For data see Supplementary Information Table S3, column 5

## Discussion

In the fourth round of the REACT-2 study of SARS-CoV-2 antibody prevalence in England during late October to early November 2020, we observed a significant increase in the proportion of the population with detectable antibodies compared with August and September 2020, especially in younger people, students, and in the Northern Regions. This is consistent with evidence that SARS-CoV-2 infection rates have been rising since late summer, with highest prevalence and rates of increase in the 18 to 24 year olds and in the North of England.[5,14]

As previously reported with respect to the first wave, we found increased prevalence among healthcare and care home workers, people of Black and Asian ethnicities, and those living in more deprived areas, as well as in London and among younger people (ages 18 to 24 years).[6] We also found that essential workers such as those in education, public transport and other public-facing roles had higher antibody prevalence than non-essential workers. These results indicate that variation and inequities in risk of infection noted in the first wave are persisting into the second wave. Antibody prevalence and rate of increase were particularly high in students, most likely reflecting the increased social mixing in this group when the UK Government advised universities to allow students to return to campus in Autumn 2020.

Our study has limitations. Participation bias may have been introduced from the lower response from ethnic minority groups and people in more deprived areas. Therefore, it is possible that our study is not fully representative of the population of England. However, we attempted to minimise this by correcting for differential response rates and differences in population characteristics introduced by our sampling strategy. An important limitation was the exclusion of children for regulatory reasons as the LFIA was approved for research use in adults only. Given that the highest antibody prevalence and increases were observed in students and younger adults, not sampling children, in particular those of secondary school-age, means that we were unable to better understand infection risk factors in the youngest age groups. We used at-home self-adminitered LFIAs which despite being a cost-effective solution to conducting large-scale surveillance studies are generally less sensitive than laboratory assays.[9] However, our comprehensive laboratory evaluation and usability studies of the selected LFIA found it’s performance to be acceptable, in terms of sensitivity and specificity in comparision to a “gold standard” ELISA test,[9] and feasible to use at-home by members of the public, in terms of usability of the LFIA kit and achieving a valid test result.[15]

In summary, although we observed a significant decline in the SARS-CoV-2 antibody prevalence in the adult population in England from June through to September 2020 after the first peak of infections, the antibody prevalence is increasing into mid-Autumn 2020. This pattern is consistent with declining population antibody levels to SARS-CoV-2 over time in the presence of low levels of transmission,[7] as seen in England in summer 2020;[14] followed by levels rising again after increases in transmission and infection rates, as seen in England from the end of August 2020.[5] By late October a large proportion of the population remained susceptible to SARS-CoV-2 infection in England based on naturally acquired immunity from the first and early second wave. To limit the number of hospitalisation and deaths in a second wave, until effective vaccines are widely available, we must continue to support the public to comply with established non pharmaceutical interventions (NPIs), including social distancing, frequent hand-washing, and face covers.

## Data Availability

Summary tabular data are provided with this paper.

https://www.imperial.ac.uk/medicine/research-and-impact/groups/react-study/

## Data availability

Summary tabular data are provided with this paper.

## Declaration of interests

We declare no competing interests.

## Funding

This work was funded by the Department of Health and Social Care in England. The content of this manuscript and decision to submit for publication were the responsibility of the authors and the funders had no role in these decisions.

## Acknowledgements

HW is a National Institute for Health Research (NIHR) Senior Investigator and acknowledges support from NIHR Biomedical Research Centre of Imperial College NHS Trust, NIHR School of Public Health Research, NIHR Applied Research Collaborative North West London, and Wellcome Trust (UNS32973). GC is supported by an NIHR Professorship. WSB is the Action Medical Research Professor, AD is an NIHR senior investigator and DA and PE are Emeritus NIHR Senior Investigators. SR and CAD acknowledge support from MRC Centre for Global Infectious Disease Analysis, National Institute for Health Research (NIHR) Health Protection Research Unit (HPRU), Wellcome Trust (200861/Z/16/Z, 200187/Z/15/Z), and Centres for Disease Control and Prevention (US, U01CK0005-01-02). PE is Director of the MRC Centre for Environment and Health (MR/L01341X/1, MR/S019669/1). PE acknowledges support from the NIHR Imperial Biomedical Research Centre and the NIHR HPRUs in Chemical and Radiation Threats and Hazards and in Environmental Exposures and Health, the British Heart Foundation Centre for Research Excellence at Imperial College London (RE/18/4/34215), Health Data Research UK (HDR UK) and the UK Dementia Research Institute at Imperial (MC_PC_17114). We thank The Huo Family Foundation for their support of our work on COVID-19.

We thank key collaborators on this work -- Ipsos MORI: Stephen Finlay, John Kennedy, Kevin Pickering, Duncan Peskett, Sam Clemens and Kelly Beaver; Institute of Global Health Innovation at Imperial College London: Gianluca Fontana, Dr Hutan Ashrafian, Sutha Satkunarajah, Didi Thompson and Lenny Naar; the Imperial Patient Experience Research Centre and the REACT Public Advisory Panel; NHS Digital for access to the NHS Register.

## Notes

### Competing Interest Statement

The authors have declared no competing interest.

### Author Declarations

We obtained research ethics approval from the South Central-Berkshire B Research Ethics Committee (IRAS ID: 283787), and Medicines and Healthcare products Regulatory Agency approval for use of the LFIA for research purposes only.

