## Supplementary Material for "Increasing SARS-CoV-2 antibody prevalence in England at the start of the second wave: REACT-2 Round 4 cross-sectional study in 160,000 adults"

**REACT-2 Round 4: Supplementary Information**

### Table S1: REACT-2 Round 4: Participation rates

|  | **Round 4 (27 Oct – 10 Nov)** | **Response (of all invitations)** | **Response (of all registered)*** |
| --- | --- | --- | --- |
| Invitation letters sent | 565000 |  |  |
| Registered* | 200721 | 36% |  |
| Requested test kit | 193611 | 34% | 96% |
| Completed survey | 169927 | 30% | 85% |
| Attempted test | 166636 | 29% | 83% |
| Completed test | 164213 | 29% | 82% |
| Valid test | 161537 | 29% | 80% |

*registration closed when > 200,000 had signed up

*Table S2: React-2 Round 4: SARS CoV-2 antibody prevalence by sociodemographic and clinical covariates (27 Oct – 10 Nov 2020, n=161,537)*

| **Category** | **Total antibody positive** | **Total tests (with valid results)** | **Crude prevalence (%) [95% CI]** | **Prevalence adjusted for test (%) [95% CI]** | **Weighted^1^ prevalence (%) [95% CI]** |
| --- | --- | --- | --- | --- | --- |
| **England** |  |  |  |  |  |
| All | 8431 | 161537 | 5.22 [5.11-5.33] | 4.60 [4.47-4.73] | 5.56 [5.43-5.71] |
| **Sex** |  |  |  |  |  |
| Male | 3598 | 70727 | 5.09 [4.93-5.25] | 4.44 [4.25-4.64] | 5.51 [5.31-5.71] |
| Female | 4833 | 90810 | 5.32 [5.18-5.47] | 4.73 [4.55-4.90] | 5.62 [5.42-5.82] |
| **Age** |  |  |  |  |  |
| 18-24 | 851 | 9539 | 8.92 [8.37-9.51] | 9.06 [8.39-9.77] | 9.85 [9.33-10.38] |
| 25-34 | 1287 | 21049 | 6.11 [5.80-6.45] | 5.68 [5.30-6.08] | 6.60 [6.25-6.96] |
| 35-44 | 1365 | 26884 | 5.08 [4.82-5.35] | 4.43 [4.12-4.75] | 5.28 [4.94-5.63] |
| 45-54 | 1812 | 32799 | 5.52 [5.28-5.78] | 4.97 [4.68-5.27] | 5.48 [5.16-5.82] |
| 55-64 | 1767 | 33296 | 5.31 [5.07-5.55] | 4.71 [4.42-5.00] | 5.53 [5.17-5.89] |
| 65-74 | 981 | 26465 | 3.71 [3.49-3.94] | 2.78 [2.51-3.06] | 3.35 [3.03-3.70] |
| 75+ | 368 | 11505 | 3.20 [2.89-3.54] | 2.17 [1.80-2.57] | 2.63 [2.30-2.98] |
| **Ethnicity** |  |  |  |  |  |
| White | 7450 | 150527 | 4.95 [4.84-5.06] | 4.28 [4.15-4.41] | 4.80 [4.66-4.94] |
| Mixed | 140 | 1959 | 7.15 [6.09-8.37] | 6.92 [5.65-8.40] | 7.18 [5.83-8.75] |
| Asian^2^ | 527 | 5425 | 9.71 [8.95-10.53] | 10.02 [9.10-11.00] | 11.65 [10.96-12.38] |
| Black^3^ | 119 | 1180 | 10.08 [8.49-11.93] | 10.46 [8.55-12.69] | 11.80 [10.66-13.04] |
| Other | 132 | 1315 | 10.04 [8.53-11.78] | 10.41 [8.59-12.51] | 13.36 [11.54-15.40] |
| **Region** |  |  |  |  |  |
| North East | 359 | 6024 | 5.96 [5.39-6.59] | 5.49 [4.81-6.25] | 5.55 [4.95-6.21] |
| North West | 1304 | 19142 | 6.81 [6.46-7.18] | 6.52 [6.10-6.96] | 7.67 [7.24-8.12] |
| Yorkshire and Humber | 588 | 11078 | 5.31 [4.91-5.74] | 4.71 [4.22-5.23] | 6.33 [5.88-6.81] |
| East Midlands | 942 | 20747 | 4.54 [4.27-4.83] | 3.78 [3.45-4.14] | 4.16 [3.75-4.60] |
| West Midlands | 879 | 15261 | 5.76 [5.40-6.14] | 5.25 [4.82-5.71] | 6.02 [5.59-6.48] |
| East of England | 1061 | 23004 | 4.61 [4.35-4.89] | 3.87 [3.55-4.21] | 4.20 [3.83-4.59] |
| London | 1384 | 15544 | 8.90 [8.47-9.36] | 9.04 [8.51-9.59] | 9.54 [9.10-9.98] |
| South East | 1407 | 35170 | 4.00 [3.80-4.21] | 3.13 [2.89-3.39] | 3.25 [2.97-3.54] |
| South West | 507 | 15567 | 3.26 [2.99-3.55] | 2.24 [1.91-2.59] | 2.29 [1.97-2.63] |
| **IMD quintile^4^** |  |  |  |  |  |
| Most deprived: 1 | 999 | 15727 | 6.35 [5.98-6.74] | 5.97 [5.52-6.44] | 7.22 [6.87-7.57] |
| 2 | 1332 | 25492 | 5.23 [4.96-5.51] | 4.61 [4.29-4.95] | 5.62 [5.32-5.94] |
| 3 | 1815 | 34784 | 5.22 [4.99-5.46] | 4.60 [4.32-4.89] | 5.49 [5.18-5.80] |
| 4 | 2007 | 40259 | 4.99 [4.78-5.20] | 4.32 [4.07-4.58] | 4.88 [4.59-5.19] |
| Least deprived: 5 | 2278 | 45275 | 5.03 [4.83-5.24] | 4.38 [4.14-4.62] | 4.66 [4.37-4.97] |
| **Highest educational level reached** |  |  |  |  |  |
| No qualification | 643 | 13923 | 4.62 [4.28-4.98] | 3.88 [3.47-4.31] |  |
| Other | 495 | 10309 | 4.80 [4.41-5.23] | 4.10 [3.62-4.62] |  |
| GCSE | 1632 | 34147 | 4.78 [4.56-5.01] | 4.07 [3.80-4.35] |  |
| Post-GCSE qualification | 2540 | 44526 | 5.70 [5.49-5.92] | 5.19 [4.93-5.45] |  |
| Degree or above | 3080 | 57487 | 5.36 [5.18-5.54] | 4.77 [4.55-4.99] |  |
| **Gross household income** |  |  |  |  |  |
| £0-14,999 | 459 | 10363 | 4.43 [4.05-4.84] | 3.65 [3.19-4.15] |  |
| £15,000-49,999 | 2316 | 46899 | 4.94 [4.75-5.14] | 4.26 [4.03-4.50] |  |
| £50,000-149,999 | 2381 | 41196 | 5.78 [5.56-6.01] | 5.28 [5.01-5.55] |  |
| >£150,000 | 377 | 5272 | 7.15 [6.49-7.88] | 6.93 [6.13-7.81] |  |
| **Employment** |  |  |  |  |  |
| Healthcare (patient-facing) | 589 | 6039 | 9.75 [9.03-10.53] | 10.06 [9.19-11.00] |  |
| Healthcare (other) | 133 | 1957 | 6.80 [5.76-8.00] | 6.50 [5.26-7.95] |  |
| Care home (client-facing) | 94 | 853 | 11.02 [9.09-13.30] | 11.59 [9.27-14.34] |  |
| Care home (other) | 21 | 233 | 9.01 [5.97-13.38] | 9.17 [5.51-14.44] |  |
| Other essential worker**^5^** | 1799 | 31787 | 5.66 [5.41-5.92] | 5.13 [4.83-5.44] |  |
| Other worker | 3197 | 59632 | 5.36 [5.18-5.54] | 4.77 [4.56-4.99] |  |
| Not in employment | 2500 | 59546 | 4.20 [4.04-4.36] | 3.37 [3.18-3.57] |  |
| **Employment (detailed)** |  |  |  |  |  |
| Full time | 4055 | 66389 | 6.11 [5.93-6.29] | 5.67 [5.46-5.89] |  |
| Part time | 1065 | 19612 | 5.43 [5.12-5.76] | 4.86 [4.48-5.25] |  |
| Self-employed | 811 | 15990 | 5.07 [4.74-5.42] | 4.42 [4.03-4.85] |  |
| Govt supported training | 6 | 61 | 9.84 [4.59-19.84] | 10.16 [3.84-22.22] |  |
| Unemployed | 187 | 3480 | 5.37 [4.67-6.17] | 4.79 [3.94-5.75] |  |
| Retired | 1269 | 37125 | 3.42 [3.24-3.61] | 2.43 [2.21-2.66] |  |
| Student | 337 | 2950 | 11.42 [10.33-12.62] | 12.08 [10.75-13.52] |  |
| Looking after home/family | 246 | 5949 | 4.14 [3.66-4.67] | 3.30 [2.72-3.94] |  |
| Sick/disabled | 102 | 2848 | 3.58 [2.96-4.33] | 2.63 [1.88-3.53] |  |
| Other | 131 | 2672 | 4.90 [4.15-5.79] | 4.22 [3.31-5.29] |  |
| Prefer not to say | 222 | 4461 | 4.98 [4.38-5.65] | 4.31 [3.59-5.13] |  |
| **Employment (sector)** |  |  |  |  |  |
| Home delivery | 102 | 1805 | 5.65 [4.68-6.81] | 5.12 [3.95-6.52] |  |
| Retail | 343 | 6406 | 5.35 [4.83-5.93] | 4.76 [4.13-5.46] |  |
| Emergency services, prisons, coastguard | 100 | 1481 | 6.75 [5.58-8.15] | 6.45 [5.04-8.13] |  |
| Public transport or taxi | 84 | 1161 | 7.24 [5.88-8.87] | 7.03 [5.40-9.00] |  |
| Teacher or childcare | 704 | 11448 | 6.15 [5.72-6.60] | 5.72 [5.21-6.27] |  |
| Armed forces | 4 | 143 | 2.80 [1.09-6.97] | 1.68 [0.00-6.71] |  |
| Other | 2243 | 39062 | 5.74 [5.52-5.98] | 5.23 [4.96-5.51] |  |
| Not working outside home | 1673 | 33005 | 5.07 [4.84-5.31] | 4.42 [4.14-4.71] |  |
| **Household size** |  |  |  |  |  |
| 1 | 1141 | 24889 | 4.58 [4.33-4.85] | 3.84 [3.53-4.16] |  |
| 2 | 2822 | 62314 | 4.53 [4.37-4.69] | 3.77 [3.58-3.97] |  |
| 3 | 1794 | 32057 | 5.60 [5.35-5.85] | 5.06 [4.76-5.37] |  |
| 4 | 1724 | 29491 | 5.85 [5.58-6.12] | 5.36 [5.04-5.69] |  |
| 5 | 595 | 8938 | 6.66 [6.16-7.19] | 6.33 [5.73-6.98] |  |
| 6 | 209 | 2537 | 8.24 [7.23-9.37] | 8.24 [7.02-9.60] |  |
| 7+ | 146 | 1311 | 11.14 [9.55-12.95] | 11.73 [9.81-13.92] | 13.45 [11.82-15.26] |
| **One or more children (under 18) in household** |  |  |  |  |  |
| No | 5699 | 111275 | 5.12 [4.99-5.25] | 4.48 [4.33-4.64] | 5.31 [5.14-5.48] |
| Yes | 2732 | 50262 | 5.44 [5.24-5.64] | 4.86 [4.63-5.10] |  |
| **History of COVID-19 symptoms** |  |  |  |  |  |
| No symptoms | 188 | 1247 | 15.08 [13.20-17.17] | 16.48 [14.21-19.00] |  |
| Mild symptoms | 1422 | 8506 | 16.72 [15.94-17.53] | 18.45 [17.52-19.43] |  |
| Moderate symptoms | 2720 | 14019 | 19.40 [18.76-20.07] | 21.69 [20.91-22.49] |  |
| Severe symtoms | 1328 | 5841 | 22.74 [21.68-23.83] | 25.71 [24.43-27.02] | 26.79 [25.51-28.11] |
| **BMI^6^** |  |  |  |  |  |
| Underweight (<18.5) | 89 | 1975 | 4.51 [3.68-5.51] | 3.74 [2.74-4.96] |  |
| Normal (18.5-24.9) | 2965 | 59112 | 5.02 [4.84-5.19] | 4.36 [4.15-4.57] |  |
| Overweight (25-29.9) | 2664 | 51230 | 5.20 [5.01-5.40] | 4.58 [4.35-4.81] |  |
| Obese (>=30) | 1816 | 30311 | 5.99 [5.73-6.26] | 5.53 [5.22-5.86] |  |
| **Current smoker** |  |  |  |  |  |
| Yes | 580 | 15884 | 3.65 [3.37-3.95] | 2.71 [2.37-3.08] |  |
| No | 7777 | 144378 | 5.39 [5.27-5.50] | 4.80 [4.66-4.94] |  |
| **Population density quintile** |  |  |  |  |  |
| 1 | 1161 | 31036 | 3.74 [3.54-3.96] | 2.82 [2.57-3.08] |  |
| 2 | 1536 | 30893 | 4.97 [4.74-5.22] | 4.30 [4.02-4.60] |  |
| 3 | 1531 | 30787 | 4.97 [4.74-5.22] | 4.30 [4.02-4.60] |  |
| 4 | 1560 | 30304 | 5.15 [4.90-5.40] | 4.52 [4.22-4.82] |  |
| 5 | 2643 | 38517 | 6.86 [6.61-7.12] | 6.58 [6.28-6.89] | 7.64 [7.38-7.91] |
| **History of COVID-19^7^** |  |  |  |  |  |
| Positive test | 1094 | 1765 | 61.98 [59.69-64.22] | 72.99 [70.23-75.69] |  |
| Suspected by doctor | 482 | 1850 | 26.05 [24.11-28.10] | 29.70 [27.36-32.17] |  |
| Suspected by respondent | 3950 | 25777 | 15.32 [14.89-15.77] | 16.78 [16.25-17.31] |  |
| No | 2773 | 131924 | 2.10 [2.03-2.18] | 0.85 [0.75-0.94] |  |
| **Time since positive PCR test** |  |  |  |  |  |
| <30 days ago | 431 | 798 | 54.01 [50.54-57.44] | 63.39 [59.21-67.52] |  |
| 30-60 days ago | 168 | 218 | 77.06 [71.04-82.15] | 91.16 [83.91-97.29] |  |
| 61-90 days ago | 38 | 66 | 57.58 [45.56-68.76] | 67.68 [53.21-81.15] |  |
| 90+ days ago | 457 | 683 | 66.91 [63.30-70.34] | 78.93 [74.57-83.06] |  |
| **Time since COVID-19 symptom onset** |  |  |  |  |  |
| <30 days ago | 525 | 1864 | 28.17 [26.17-30.25] | 32.25 [29.84-34.76] |  |
| 30-60 days ago | 317 | 1134 | 27.95 [25.42-30.64] | 31.99 [28.94-35.23] |  |
| 61-90 days ago | 48 | 374 | 12.83 [9.82-16.61] | 13.78 [10.14-18.32] |  |
| 91-120 days ago | 27 | 227 | 11.89 [8.30-16.75] | 12.64 [8.32-18.50] |  |
| 121-150 days ago | 36 | 257 | 14.01 [10.29-18.78] | 15.19 [10.71-20.94] |  |
| >150 days ago | 4331 | 20642 | 20.98 [20.43-21.54] | 23.59 [22.93-24.27] |  |
| **Contact with COVID-19 case^8^** |  |  |  |  |  |
| Yes, with confirmed case | 1890 | 12871 | 14.68 [14.08-15.31] | 16.01 [15.28-16.75] |  |
| Yes, with suspected case | 1111 | 6920 | 16.05 [15.21-16.94] | 17.66 [16.64-18.72] |  |
| No | 5430 | 141746 | 3.83 [3.73-3.93] | 2.93 [2.81-3.05] |  |
| **Number of existing health conditions^9^** |  |  |  |  |  |
| >1 | 1216 | 25468 | 4.77 [4.52-5.04] | 4.07 [3.76-4.39] |  |
| 1 | 2022 | 41362 | 4.89 [4.68-5.10] | 4.20 [3.96-4.46] |  |
| 0 | 5193 | 94707 | 5.48 [5.34-5.63] | 4.92 [4.75-5.10] |  |
| **Care home resident** |  |  |  |  |  |
| Yes | 25 | 313 | 7.99 [5.47-11.52] | 7.94 [4.90-12.20] |  |
| No | 8406 | 161224 | 5.21 [5.11-5.32] | 4.60 [4.47-4.73] |  |
| **Reported symptoms of previous COVID-19 case^10^** |  |  |  |  |  |
| No symptoms | 2982 | 133318 | 2.24 [2.16-2.32] | 1.01 [0.91-1.11] |  |
| Atypical symptoms only | 571 | 5011 | 11.39 [10.54-12.30] | 12.04 [11.02-13.14] |  |
| Screening symptoms | 4878 | 23208 | 21.02 [20.50-21.55] | 23.64 [23.01-24.27] |  |
| **Number of children (under 18) in household** |  |  |  |  |  |
| 0 | 5699 | 111275 | 5.12 [4.99-5.25] | 4.48 [4.33-4.64] |  |
| 1 | 1238 | 22282 | 5.56 [5.26-5.86] | 5.01 [4.65-5.38] |  |
| 2 | 1127 | 21645 | 5.21 [4.92-5.51] | 4.59 [4.24-4.95] |  |
| >2 | 367 | 6335 | 5.79 [5.24-6.40] | 5.29 [4.63-6.02] | 6.58 [5.88-7.34] |
| **COVID outcome severity** |  |  |  |  |  |
| No treatment | 3488 | 17809 | 19.59 [19.01-20.17] | 21.91 [21.22-22.62] |  |
| Sought medical care | 1589 | 8636 | 18.40 [17.60-19.23] | 20.48 [19.51-21.48] |  |
| Hospital admission | 107 | 369 | 29.00 [24.60-33.82] | 33.25 [27.96-39.07] |  |
| ICU | 20 | 49 | 40.82 [28.22-54.75] | 47.49 [32.31-64.28] |  |
| **Number of children under 5 in household** |  |  |  |  |  |
| 0 | 7734 | 147006 | 5.26 [5.15-5.38] | 4.65 [4.52-4.79] |  |
| 1 | 589 | 12143 | 4.85 [4.48-5.25] | 4.16 [3.71-4.64] |  |
| 2 | 104 | 2332 | 4.46 [3.69-5.38] | 3.69 [2.76-4.79] |  |
| >2 | 4 | 56 | 7.14 [2.81-16.98] | 6.92 [1.70-18.77] |  |
| **Number of children aged 5 to 10 in household** |  |  |  |  |  |
| 0 | 7469 | 142778 | 5.23 [5.12-5.35] | 4.62 [4.48-4.76] |  |
| 1 | 738 | 14558 | 5.07 [4.72-5.44] | 4.42 [4.01-4.86] |  |
| 2 | 205 | 3947 | 5.19 [4.54-5.93] | 4.57 [3.79-5.46] |  |
| >2 | 19 | 254 | 7.48 [4.84-11.39] | 7.33 [4.15-12.03] |  |
| **Number of children aged 11 to 17 in household** |  |  |  |  |  |
| 0 | 7052 | 138160 | 5.10 [4.99-5.22] | 4.46 [4.32-4.60] |  |
| 1 | 969 | 16593 | 5.84 [5.49-6.21] | 5.35 [4.93-5.79] |  |
| 2 | 372 | 6145 | 6.05 [5.48-6.68] | 5.61 [4.92-6.36] |  |
| >2 | 38 | 639 | 5.95 [4.36-8.06] | 5.48 [3.57-8.02] |  |
| **Ethnicity (granular)** |  |  |  |  |  |
| White British | 6856 | 141124 | 4.86 [4.75-4.97] | 4.17 [4.03-4.30] |  |
| White Irish | 108 | 1519 | 7.11 [5.92-8.51] | 6.88 [5.45-8.57] |  |
| White gypsy or Irish traveller | 1 | 35 | 2.86 [0.51-14.53] | 1.76 [0.00-15.82] |  |
| Any other white background | 485 | 7849 | 6.18 [5.67-6.73] | 5.76 [5.14-6.43] |  |
| Mixed white / black Carribean | 35 | 427 | 8.20 [5.95-11.19] | 8.19 [5.48-11.79] |  |
| mixed white / black African | 12 | 179 | 6.70 [3.88-11.35] | 6.39 [2.98-11.99] |  |
| Mixed white and Asian | 51 | 682 | 7.48 [5.73-9.70] | 7.32 [5.22-10.00] |  |
| Any other mixed / multiple ethnic background | 42 | 671 | 6.26 [4.66-8.35] | 5.85 [3.93-8.38] |  |
| Indian | 260 | 2896 | 8.98 [7.99-10.07] | 9.13 [7.94-10.45] |  |
| Pakistani | 89 | 686 | 12.97 [10.66-15.70] | 13.94 [11.16-17.22] |  |
| Bangladeshi | 41 | 294 | 13.95 [10.45-18.37] | 15.12 [10.90-20.45] |  |
| Chinese | 38 | 608 | 6.25 [4.59-8.46] | 5.84 [3.84-8.51] |  |
| Other Asian | 99 | 941 | 10.52 [8.72-12.64] | 10.99 [8.82-13.55] |  |
| African | 84 | 668 | 12.57 [10.27-15.31] | 13.46 [10.69-16.75] |  |
| Caribbean | 29 | 414 | 7.00 [4.92-9.88] | 6.75 [4.24-10.22] |  |
| Other black / African / Caribbean background | 6 | 98 | 6.12 [2.84-12.72] | 5.69 [1.73-13.64] |  |
| Arab | 24 | 232 | 10.34 [7.05-14.93] | 10.78 [6.81-16.30] |  |
| Any other ethnic group | 108 | 1083 | 9.97 [8.33-11.90] | 10.33 [8.35-12.65] |  |
| Prefer not to say | 63 | 1130 | 5.58 [4.38-7.07] | 5.03 [3.59-6.83] |  |
| **Employment (broad)** |  |  |  |  |  |
| Healthcare | 722 | 7996 | 9.03 [8.42-9.68] | 9.19 [8.46-9.97] |  |
| Care home | 115 | 1086 | 10.59 [8.90-12.56] | 11.07 [9.03-13.45] |  |
| Other essential worker**^5^** | 1799 | 31787 | 5.66 [5.41-5.92] | 5.13 [4.83-5.44] |  |
| Other worker | 3197 | 59632 | 5.36 [5.18-5.54] | 4.77 [4.56-4.99] |  |
| Not in employment | 2500 | 59546 | 4.20 [4.04-4.36] | 3.37 [3.18-3.57] |  |
| **Age group (broad)** |  |  |  |  |  |
| 18-24 | 851 | 9539 | 8.92 [8.37-9.51] | 9.06 [8.39-9.77] |  |
| 25-44 | 2652 | 47933 | 5.53 [5.33-5.74] | 4.98 [4.74-5.23] |  |
| 45-64 | 3579 | 66095 | 5.41 [5.24-5.59] | 4.84 [4.63-5.05] |  |
| 65+ | 1349 | 37970 | 3.55 [3.37-3.74] | 2.59 [2.38-2.82] |  |

1 All estimates of prevalence adjusted for imperfect test sensitivity and specificity (see text for details). Responses have been re-weighted to account for sample design and for variation in response rate (age, gender, ethnicity and deprivation) in final column to be representative of the England population (18+); 2 Asian / Asian British; 3 Black / African / Caribbean / Black British; 4 Based on Index of Multiple Deprivation (2019) at lower super output area. 5 List of essential workers from UK government <https://www.gov.uk/guidance/coronavirus-covid-19-getting-tested#essential-workers> 6 Body Mass Index (BMI) formula: weight (kg) / (height [m])^2^. 7 COVID History is self-reported, based on response to the question, “Before you took this antibody test, did you think you had had COVID-19?” with response options of Yes, confirmed by a positive test (swab/*PCR/antigen test); Yes, suspected by a doctor but not tested; Yes, my own suspicions; No. 8 Self reported contact with a case of COVID19.9 Pre-existing health conditions included: organ transplant recipient, diabetes (type I or II), heart disease or heart problems, hypertension, overweight, stroke, kidney disease, liver disease, anemia, asthma, other lung condition, cancer, condition affecting the brain and nerves, weakened immune system/reduced ability to deal with infections, depression, anxiety, psychiatric disorder, none of these. 10 Symptom category is constructed from responses about self-reported specific symptoms. These were grouped into those reporting one or more “screening symptoms”nbased on recommendations for having a SARS-CoV-2 test (new persistent cough, fever, loss of sense of smell or taste), or atypical (any other symptom(s)), or none.

### Table S3: REACT-2 Round 4: Logistic regression: odds ratios for antibody positivity

Unadjusted and adjusted odds ratios [95% confidence intervals]. Unadjusted odds ratios were obtained from univariable logistic regression for the covariate of interest. Adjusted odds ratios were obtained by performing multivariable logistic regression for the covariate of interest with age and gender (column 3) or age, gender, and region (column 4). Column 5 gives adjusted odds ratios from multivariable logistic regression adjusting for age, sex, ethnicity, region, deprivation, population density, household size and keyworker status.

| **Covariate** | **Unadjusted** | **Adjusted for Age and Gender** | **Adjusted for Age, Gender, Region** | **Multivariable Adjusted** |
| --- | --- | --- | --- | --- |
| **Sex** |  |  |  |  |
| Male | Reference | Reference | reference | reference |
| Female | 1.05 [1.00,1.10] | 1.02 [0.98,1.07] | 1.03 [0.98,1.07] | 1.01 [0.96,1.06] |
| **Age** |  |  |  |  |
| 18-24 | 1.83 [1.68,2.00] | 1.83 [1.67,2.00] | 1.86 [1.70,2.03] | 1.93 [1.76,2.12] |
| 25-34 | 1.22 [1.13,1.32] | 1.22 [1.12,1.32] | 1.18 [1.09,1.28] | 1.23 [1.14,1.33] |
| 35-44 | Reference | Reference | reference | reference |
| 45-54 | 1.09 [1.02,1.18] | 1.09 [1.02,1.18] | 1.13 [1.05,1.21] | 1.17 [1.08,1.25] |
| 55-64 | 1.05 [0.97,1.13] | 1.05 [0.98,1.13] | 1.09 [1.02,1.17] | 1.25 [1.15,1.35] |
| 65-74 | 0.72 [0.66,0.78] | 0.72 [0.66,0.78] | 0.76 [0.70,0.83] | 1.01 [0.91,1.11] |
| 75+ | 0.62 [0.55,0.69] | 0.62 [0.55,0.70] | 0.66 [0.58,0.74] | 0.91 [0.80,1.04] |
| **Ethnicity** |  |  |  |  |
| White | Reference | Reference | reference | reference |
| Mixed | 1.48 [1.24,1.76] | 1.29 [1.09,1.54] | 1.19 [1.00,1.42] | 1.19 [1.00,1.42] |
| Asian | 2.07 [1.88,2.27] | 1.94 [1.77,2.13] | 1.68 [1.52,1.85] | 1.56 [1.42,1.72] |
| Black | 2.15 [1.78,2.61] | 2.04 [1.69,2.48] | 1.71 [1.40,2.07] | 1.55 [1.27,1.89] |
| Other | 2.14 [1.79,2.57] | 2.09 [1.74,2.50] | 1.75 [1.46,2.11] | 1.71 [1.42,2.06] |
| **Region** |  |  |  |  |
| North East | 1.52 [1.35,1.71] | 1.51 [1.34,1.70] | 1.51 [1.34,1.70] | 1.53 [1.36,1.73] |
| North West | 1.75 [1.62,1.90] | 1.75 [1.62,1.89] | 1.75 [1.62,1.89] | 1.76 [1.62,1.90] |
| Yorkshire and The Humber | 1.35 [1.22,1.48] | 1.34 [1.21,1.48] | 1.34 [1.21,1.48] | 1.36 [1.23,1.50] |
| East Midlands | 1.14 [1.05,1.24] | 1.14 [1.04,1.24] | 1.14 [1.04,1.24] | 1.15 [1.06,1.25] |
| West Midlands | 1.47 [1.35,1.60] | 1.46 [1.34,1.59] | 1.46 [1.34,1.59] | 1.46 [1.33,1.59] |
| East of England | 1.16 [1.07,1.26] | 1.15 [1.06,1.25] | 1.15 [1.06,1.25] | 1.15 [1.06,1.25] |
| London | 2.35 [2.17,2.53] | 2.25 [2.09,2.44] | 2.25 [2.09,2.44] | 2.14 [1.98,2.32] |
| South East | Reference | Reference | reference | reference |
| South West | 0.81 [0.73,0.90] | 0.82 [0.74,0.90] | 0.82 [0.74,0.90] | 0.82 [0.74,0.91] |
| **IMD quintile** |  |  |  |  |
| Most deprived: 1 | 1.28 [1.19,1.38] | 1.21 [1.12,1.31] | 1.04 [0.96,1.12] | 1.01 [0.93,1.09] |
| 2 | 1.04 [0.97,1.12] | 1.01 [0.94,1.08] | 0.89 [0.83,0.96] | 0.88 [0.82,0.95] |
| 3 | 1.04 [0.98,1.11] | 1.02 [0.96,1.09] | 0.97 [0.91,1.03] | 0.96 [0.90,1.03] |
| 4 | 0.99 [0.93,1.05] | 0.98 [0.92,1.05] | 0.94 [0.88,1.00] | 0.94 [0.88,1.00] |
| Least deprived: 5 | Reference | Reference | reference | reference |
| **Highest educational level reached** |  |  |  |  |
| No qualification | Reference | Reference | reference | reference |
| Other | 1.04 [0.92,1.17] | 0.99 [0.87,1.11] | 0.98 [0.87,1.11] | 0.95 [0.84,1.07] |
| GCSE | 1.04 [0.94,1.14] | 0.85 [0.77,0.94] | 0.88 [0.80,0.97] | 0.90 [0.81,0.99] |
| Post-GCSE qualification | 1.25 [1.14,1.37] | 0.95 [0.87,1.05] | 0.98 [0.89,1.08] | 0.97 [0.88,1.06] |
| Degree or above | 1.17 [1.07,1.28] | 0.92 [0.84,1.01] | 0.89 [0.81,0.98] | 0.86 [0.78,0.94] |
| **Gross household income** |  |  |  |  |
| £0-14,999 | 0.89 [0.81,0.99] | 0.91 [0.82,1.01] | 0.89 [0.80,0.98] | 0.92 [0.82,1.02] |
| £15,000-49,999 | Reference | Reference | reference | reference |
| £50,000-149,999 | 1.18 [1.11,1.25] | 1.11 [1.04,1.18] | 1.09 [1.03,1.16] | 1.09 [1.02,1.16] |
| >£150,000 | 1.48 [1.32,1.66] | 1.39 [1.24,1.55] | 1.26 [1.12,1.42] | 1.26 [1.11,1.42] |
| **Employment** |  |  |  |  |
| Healthcare (patient-facing) | 1.91 [1.74,2.09] | 1.89 [1.72,2.07] | 1.93 [1.76,2.12] | 1.86 [1.69,2.04] |
| Healthcare (other) | 1.29 [1.08,1.54] | 1.30 [1.09,1.56] | 1.33 [1.11,1.59] | 1.32 [1.10,1.58] |
| Care home (client-facing) | 2.19 [1.76,2.72] | 2.17 [1.74,2.70] | 2.30 [1.85,2.87] | 2.19 [1.75,2.74] |
| Care home (other) | 1.75 [1.12,2.74] | 1.77 [1.13,2.78] | 1.88 [1.20,2.95] | 1.88 [1.20,2.96] |
| Other essential worker | 1.06 [1.00,1.12] | 1.05 [0.99,1.11] | 1.08 [1.02,1.15] | 1.07 [1.01,1.14] |
| Other worker | Reference | Reference | reference | reference |
| Not in employment | 0.77 [0.73,0.82] | 0.89 [0.84,0.95] | 0.89 [0.84,0.95] | 0.86 [0.81,0.92] |
| **Employment (detailed)** |  |  |  |  |
| Full time | Reference | Reference | reference | reference |
| Part time | 0.88 [0.82,0.95] | 0.87 [0.81,0.93] | 0.90 [0.83,0.96] | 0.86 [0.80,0.93] |
| Self-employed | 0.82 [0.76,0.89] | 0.85 [0.79,0.92] | 0.86 [0.79,0.93] | 0.89 [0.82,0.96] |
| Govt supported training | 1.68 [0.72,3.90] | 1.44 [0.62,3.36] | 1.35 [0.58,3.15] | 2.29 [0.96,5.50] |
| Unemployed | 0.87 [0.75,1.01] | 0.83 [0.71,0.96] | 0.80 [0.69,0.94] | 1.37 [1.06,1.76] |
| Retired | 0.54 [0.51,0.58] | 0.62 [0.56,0.68] | 0.63 [0.57,0.69] | 1.10 [0.89,1.37] |
| Student | 1.98 [1.76,2.23] | 1.49 [1.31,1.71] | 1.47 [1.28,1.68] | 2.31 [1.81,2.94] |
| Looking after home/family | 0.66 [0.58,0.76] | 0.66 [0.58,0.76] | 0.67 [0.59,0.77] | 1.12 [0.88,1.42] |
| Sick/disabled | 0.57 [0.47,0.70] | 0.58 [0.47,0.70] | 0.57 [0.46,0.69] | 1.00 [1.00,1.00] |
| Other | 0.79 [0.66,0.95] | 0.80 [0.67,0.95] | 0.80 [0.66,0.95] | 1.39 [1.06,1.82] |
| Prefer not to say | 0.81 [0.70,0.92] | 0.82 [0.71,0.94] | 0.80 [0.70,0.92] | 1.34 [1.05,1.72] |
| **Employment (sector)** |  |  |  |  |
| Home delivery | 1.12 [0.91,1.38] | 1.12 [0.91,1.38] | 1.21 [0.98,1.48] | 1.16 [0.94,1.44] |
| Retail | 1.06 [0.94,1.19] | 1.00 [0.89,1.13] | 1.08 [0.96,1.22] | 1.07 [0.94,1.21] |
| Emergency services, prisons, coastguard | 1.36 [1.10,1.67] | 1.35 [1.10,1.67] | 1.44 [1.16,1.77] | 1.45 [1.17,1.81] |
| Public transport or taxi | 1.46 [1.16,1.83] | 1.48 [1.18,1.86] | 1.50 [1.19,1.88] | 1.47 [1.16,1.86] |
| Teacher or childcare | 1.23 [1.12,1.34] | 1.22 [1.11,1.34] | 1.27 [1.16,1.39] | 1.28 [1.15,1.42] |
| Armed forces | 0.54 [0.20,1.46] | 0.54 [0.20,1.47] | 0.62 [0.23,1.69] | 0.64 [0.24,1.73] |
| Other | 1.14 [1.07,1.22] | 1.13 [1.06,1.21] | 1.20 [1.12,1.28] | 1.18 [1.10,1.27] |
| Not working outside home | Reference | Reference | reference | reference |
| **Household size** |  |  |  |  |
| 1 | Reference | Reference | reference | reference |
| 2 | 0.99 [0.92,1.06] | 0.96 [0.90,1.03] | 0.99 [0.92,1.06] | 0.99 [0.92,1.06] |
| 3 | 1.23 [1.14,1.33] | 1.05 [0.97,1.14] | 1.09 [1.00,1.18] | 1.07 [0.98,1.16] |
| 4 | 1.29 [1.20,1.40] | 1.10 [1.01,1.19] | 1.14 [1.05,1.24] | 1.12 [1.03,1.22] |
| 5 | 1.48 [1.34,1.64] | 1.24 [1.11,1.38] | 1.28 [1.15,1.43] | 1.23 [1.11,1.38] |
| 6 | 1.87 [1.60,2.18] | 1.51 [1.29,1.77] | 1.55 [1.33,1.82] | 1.46 [1.25,1.72] |
| 7+ | 2.61 [2.17,3.13] | 2.02 [1.68,2.43] | 2.03 [1.68,2.44] | 1.94 [1.60,2.35] |
| **One or more children (under 18) in household** |  |  |  |  |
| No | Reference | Reference | reference | reference |
| Yes | 1.06 [1.02,1.12] | 0.95 [0.90,1.00] | 0.97 [0.92,1.03] | 0.80 [0.75,0.85] |
| **History of COVID-19 symptoms** |  |  |  |  |
| No symptoms | Reference | Reference | reference | reference |
| Mild symptoms | 1.13 [0.96,1.33] | 1.18 [1.00,1.39] | 1.21 [1.03,1.43] | 1.26 [1.06,1.49] |
| Moderate symptoms | 1.36 [1.15,1.59] | 1.42 [1.21,1.67] | 1.49 [1.26,1.75] | 1.53 [1.30,1.81] |
| Severe symtoms | 1.66 [1.40,1.96] | 1.74 [1.47,2.05] | 1.82 [1.53,2.15] | 1.92 [1.62,2.29] |
| **BMI** |  |  |  |  |
| Underweight (<18.5) | 0.89 [0.72,1.11] | 0.78 [0.63,0.97] | 0.76 [0.61,0.95] | 0.75 [0.60,0.93] |
| Normal (18.5-24.9) | Reference | Reference | reference | reference |
| Overweight (25-29.9) | 1.04 [0.98,1.10] | 1.10 [1.05,1.17] | 1.12 [1.06,1.18] | 1.11 [1.05,1.18] |
| Obese (>=30) | 1.21 [1.14,1.28] | 1.25 [1.17,1.32] | 1.28 [1.20,1.36] | 1.26 [1.19,1.35] |
| **Current smoker** |  |  |  |  |
| Yes | 0.67 [0.61,0.73] | 0.62 [0.57,0.67] | 0.62 [0.56,0.67] | 0.63 [0.57,0.68] |
| No | Reference | Reference | reference | reference |
| **Population density quintile** |  |  |  |  |
| 1 | Reference | Reference | reference | reference |
| 2 | 1.35 [1.25,1.46] | 1.33 [1.23,1.44] | 1.29 [1.19,1.39] | 1.28 [1.18,1.38] |
| 3 | 1.35 [1.25,1.46] | 1.32 [1.22,1.43] | 1.26 [1.16,1.36] | 1.23 [1.13,1.33] |
| 4 | 1.40 [1.29,1.51] | 1.36 [1.26,1.47] | 1.27 [1.17,1.37] | 1.25 [1.16,1.36] |
| 5 | 1.90 [1.77,2.03] | 1.79 [1.67,1.93] | 1.46 [1.35,1.57] | 1.44 [1.33,1.56] |
| **History of COVID-19** |  |  |  |  |
| Positive test | 75.94 [68.49,84.19] | 74.35 [67.02,82.49] | 71.65 [64.53,79.54] | 66.76 [59.99,74.30] |
| Suspected by doctor | 16.41 [14.69,18.33] | 16.43 [14.70,18.35] | 15.83 [14.15,17.70] | 15.64 [13.95,17.52] |
| Suspected by respondent | 8.43 [8.01,8.87] | 8.36 [7.94,8.81] | 8.18 [7.77,8.61] | 8.21 [7.79,8.65] |
| No | Reference | Reference | reference | reference |
| **Time since positive PCR test** |  |  |  |  |
| <30 days ago | Reference | Reference | reference | reference |
| 30-60 days ago | 2.86 [2.03,4.04] | 2.78 [1.97,3.94] | 2.74 [1.93,3.89] | 2.76 [1.94,3.95] |
| 61-90 days ago | 1.16 [0.70,1.92] | 1.19 [0.71,1.99] | 1.22 [0.73,2.05] | 1.23 [0.72,2.09] |
| 90+ days ago | 1.72 [1.39,2.13] | 1.83 [1.48,2.28] | 1.97 [1.57,2.46] | 2.10 [1.64,2.70] |
| **Time since COVID-19 symptom onset** |  |  |  |  |
| <30 days ago | Reference | Reference | reference | reference |
| 30-60 days ago | 0.99 [0.84,1.17] | 0.99 [0.84,1.17] | 1.00 [0.85,1.19] | 1.01 [0.85,1.19] |
| 61-90 days ago | 0.38 [0.27,0.52] | 0.38 [0.27,0.52] | 0.38 [0.28,0.52] | 0.38 [0.27,0.53] |
| 91-120 days ago | 0.34 [0.23,0.52] | 0.35 [0.23,0.53] | 0.36 [0.24,0.54] | 0.37 [0.24,0.56] |
| 121-150 days ago | 0.42 [0.29,0.60] | 0.42 [0.29,0.61] | 0.43 [0.30,0.62] | 0.42 [0.29,0.62] |
| >150 days ago | 0.68 [0.61,0.75] | 0.69 [0.62,0.77] | 0.72 [0.65,0.80] | 0.72 [0.64,0.80] |
| **Contact with COVID-19 case** |  |  |  |  |
| Yes, with confirmed case | 4.32 [4.09,4.57] | 4.05 [3.83,4.29] | 3.88 [3.66,4.11] | 3.71 [3.49,3.94] |
| Yes, with suspected case | 4.80 [4.48,5.15] | 4.57 [4.26,4.90] | 4.39 [4.09,4.71] | 4.33 [4.03,4.65] |
| No | Reference | Reference | reference | reference |
| **Number of existing health conditions** |  |  |  |  |
| >1 | 0.86 [0.81,0.92] | 0.93 [0.87,0.99] | 0.93 [0.87,0.99] | 0.95 [0.89,1.02] |
| 1 | 0.89 [0.84,0.93] | 0.94 [0.90,1.00] | 0.94 [0.90,1.00] | 0.95 [0.90,1.00] |
| 0 | Reference | Reference | reference | reference |
| **BMI (lower)** |  |  |  |  |
| Underweight (<18.5) | 0.89 [0.72,1.11] | 0.80 [0.64,1.00] | 0.79 [0.63,0.98] | 0.77 [0.61,0.96] |
| Normal (18.5-23.4) | Reference | Reference | reference | reference |
| Overweight (23.5-27.4) | 1.01 [0.95,1.07] | 1.09 [1.02,1.16] | 1.11 [1.04,1.18] | 1.09 [1.02,1.17] |
| Obese (>=27.5) | 1.15 [1.08,1.22] | 1.23 [1.16,1.32] | 1.27 [1.19,1.36] | 1.25 [1.17,1.33] |
| **Care home resident** |  |  |  |  |
| Yes | 1.58 [1.05,2.38] | 1.58 [1.05,2.38] | 1.62 [1.08,2.45] | 1.04 [0.68,1.61] |
| No | Reference | Reference | reference | reference |
| **Reported symptoms of previous COVID-19 case** |  |  |  |  |
| No symptoms | Reference | Reference | reference | reference |
| Atypical symptoms only | 5.62 [5.11,6.18] | 5.60 [5.09,6.15] | 5.38 [4.89,5.91] | 5.34 [4.85,5.88] |
| Screening symptoms | 11.63 [11.09,12.20] | 11.53 [10.98,12.11] | 11.31 [10.77,11.88] | 11.31 [10.76,11.88] |
| **Number of children (under 18) in household** |  |  |  |  |
| 0 | Reference | Reference | reference | reference |
| 1 | 1.09 [1.02,1.16] | 0.95 [0.89,1.01] | 0.96 [0.90,1.03] | 0.84 [0.78,0.90] |
| 2 | 1.02 [0.95,1.09] | 0.93 [0.86,1.00] | 0.96 [0.90,1.04] | 0.76 [0.69,0.83] |
| >2 | 1.14 [1.02,1.27] | 1.03 [0.92,1.16] | 1.07 [0.96,1.20] | 0.67 [0.58,0.77] |
| **COVID outcome severity** |  |  |  |  |
| No treatment | Reference | Reference | reference | reference |
| Sought medical care | 0.93 [0.87,0.99] | 0.93 [0.87,1.00] | 0.93 [0.87,0.99] | 0.92 [0.86,0.98] |
| Hospital admission | 1.68 [1.34,2.11] | 1.71 [1.36,2.15] | 1.74 [1.38,2.19] | 1.64 [1.30,2.09] |
| ICU | 2.83 [1.60,5.01] | 2.98 [1.68,5.28] | 2.98 [1.67,5.32] | 2.73 [1.49,5.00] |
| **Number of children under 5 in household** |  |  |  |  |
| 0 | Reference | Reference | reference | reference |
| 1 | 0.92 [0.84,1.00] | 0.83 [0.76,0.91] | 0.85 [0.78,0.93] | 0.77 [0.70,0.85] |
| 2 | 0.84 [0.69,1.02] | 0.76 [0.62,0.93] | 0.78 [0.64,0.95] | 0.64 [0.52,0.79] |
| >2 | 1.39 [0.50,3.83] | 1.22 [0.44,3.39] | 1.28 [0.46,3.56] | 1.04 [0.37,2.91] |
| **Number of children aged 5 to 10 in household** |  |  |  |  |
| 0 | Reference | Reference | reference | reference |
| 1 | 0.97 [0.90,1.05] | 0.91 [0.84,0.99] | 0.94 [0.86,1.02] | 0.85 [0.77,0.92] |
| 2 | 0.99 [0.86,1.14] | 0.95 [0.82,1.10] | 0.99 [0.85,1.14] | 0.84 [0.72,0.98] |
| >2 | 1.46 [0.92,2.34] | 1.37 [0.86,2.20] | 1.43 [0.89,2.28] | 0.98 [0.60,1.61] |
| **Number of children aged 11 to 17 in household** |  |  |  |  |
| 0 | Reference | Reference | reference | reference |
| 1 | 1.15 [1.08,1.24] | 1.05 [0.98,1.13] | 1.07 [0.99,1.15] | 0.97 [0.89,1.04] |
| 2 | 1.20 [1.08,1.33] | 1.13 [1.01,1.26] | 1.16 [1.04,1.30] | 0.99 [0.88,1.11] |
| >2 | 1.18 [0.85,1.63] | 1.09 [0.78,1.52] | 1.14 [0.82,1.58] | 0.87 [0.62,1.22] |
| **Ethnicity (granular)** |  |  |  |  |
| White British | Reference | Reference | reference | reference |
| White Irish | 1.50 [1.23,1.83] | 1.56 [1.28,1.91] | 1.41 [1.15,1.72] | 1.41 [1.15,1.72] |
| White gypsy or Irish traveller | 0.58 [0.08,4.21] | 0.55 [0.07,4.00] | 0.58 [0.08,4.26] | 0.55 [0.08,4.07] |
| Any other white background | 1.29 [1.17,1.42] | 1.24 [1.13,1.37] | 1.10 [1.00,1.22] | 1.11 [1.00,1.22] |
| Mixed white / black Carribean | 1.75 [1.24,2.47] | 1.49 [1.05,2.11] | 1.40 [0.98,1.98] | 1.22 [0.61,2.42] |
| mixed white / black African | 1.41 [0.78,2.53] | 1.24 [0.69,2.22] | 1.14 [0.63,2.05] | 1.00 [1.00,1.00] |
| Mixed white and Asian | 1.58 [1.19,2.11] | 1.38 [1.03,1.84] | 1.26 [0.95,1.69] | 1.11 [0.58,2.15] |
| Any other mixed / multiple ethnic background | 1.31 [0.96,1.79] | 1.17 [0.86,1.61] | 1.07 [0.78,1.46] | 0.94 [0.48,1.84] |
| Indian | 1.93 [1.70,2.20] | 1.85 [1.63,2.11] | 1.62 [1.42,1.85] | 1.37 [0.96,1.96] |
| Pakistani | 2.92 [2.33,3.65] | 2.64 [2.10,3.30] | 2.22 [1.77,2.79] | 1.85 [1.24,2.77] |
| Bangladeshi | 3.17 [2.28,4.42] | 2.78 [1.99,3.87] | 2.19 [1.56,3.06] | 1.77 [1.10,2.85] |
| Chinese | 1.31 [0.94,1.81] | 1.22 [0.88,1.69] | 1.06 [0.76,1.48] | 1.00 [1.00,1.00] |
| Other Asian | 2.30 [1.87,2.84] | 2.17 [1.76,2.67] | 1.88 [1.52,2.32] | 1.63 [1.10,2.42] |
| African | 2.82 [2.24,3.55] | 2.65 [2.11,3.34] | 2.22 [1.76,2.81] | 1.83 [0.77,4.33] |
| Caribbean | 1.48 [1.01,2.15] | 1.42 [0.97,2.07] | 1.16 [0.79,1.70] | 1.06 [0.43,2.64] |
| Other black / African / Caribbean background | 1.28 [0.56,2.92] | 1.21 [0.53,2.78] | 1.07 [0.47,2.45] | 1.00 [1.00,1.00] |
| Arab | 2.26 [1.48,3.45] | 2.11 [1.38,3.23] | 1.75 [1.14,2.69] | 1.00 [1.00,1.00] |
| Any other ethnic group | 2.17 [1.78,2.65] | 2.13 [1.75,2.61] | 1.80 [1.47,2.20] | 0.99 [0.62,1.59] |
| Prefer not to say | 1.16 [0.90,1.49] | 1.16 [0.90,1.50] | 1.07 [0.83,1.39] | 1.01 [0.97,1.06] |
| **Employment (broad)** |  |  |  |  |
| Healthcare | 1.75 [1.61,1.91] | 1.74 [1.60,1.90] | 1.78 [1.64,1.94] | 1.32 [1.10,1.58] |
| Care home | 2.09 [1.72,2.54] | 2.08 [1.71,2.54] | 2.21 [1.81,2.69] | 2.19 [1.75,2.74] |
| Other essential worker | 1.06 [1.00,1.12] | 1.05 [0.99,1.11] | 1.08 [1.02,1.15] | 1.07 [1.01,1.14] |
| Other worker | Reference | Reference | reference | reference |
| Not in employment | 0.77 [0.73,0.82] | 0.89 [0.84,0.95] | 0.89 [0.84,0.95] | 0.86 [0.81,0.92] |
| **Age group (broad)** |  |  |  |  |
| 18-24 | 1.67 [1.54,1.81] | 1.00 [1.00,1.00] | 1.00 [1.00,1.00] | 1.93 [1.76,2.12] |
| 25-44 | Reference | Reference | reference | reference |
| 45-64 | 0.98 [0.93,1.03] | Inf [Inf,Inf] | Inf [0.00,Inf] | 1.25 [1.15,1.35] |
| 65+ | 0.63 [0.59,0.67] | Inf [0.00,Inf] | 0.00 [0.00,Inf] | 1.00 [1.00,1.00] |

### Table S4: REACT-2 rounds 1 – 4: Change in prevalence of antibody positivity to SARS-CoV-2 using LFIA test over four rounds from June to November by key covariates

*The table below shows prevalence across the first 4 rounds in England. ^1^ All tests with IgG positive or IgG negative result (i.e. excluding invalid/ unable to read). ^2^ All adjusted for test performance (see methods), and **weighted for the following covariates: sex, age group, region, deprivation, ethnicity.*

|  | Round 1: 20 June – 13 July 2020 | | | Round 2: 31 July – 13 August 2020 | | | Round 3: 15 – 28 September 2020 | | | Round 4: 27 October – 10 November 2020 | | |  |  |  |
| --- | --- | --- | --- | --- | --- | --- | --- | --- | --- | --- | --- | --- | --- | --- | --- |
|  | Total antibody positive | Total tests^1^ | Prevalence^2^ (%) [95% CI] | Total antibody positive | Total tests^1^ | Prevalence^2^ (%) [95% CI] | Total antibody positive | Total tests^1^ | Prevalence^2^ (%) [95% CI] | Total antibody positive | Total tests^1^ | Prevalence^2^ (%) [95% CI] | Relative change in prevalence round 1 to round 2 % [95% CI] | Relative change in prevalence round 2 to round 3 % [95% CI] | Relative change in prevalence round 3 to round 4 % [95% CI] |
| **Full_cohort**** |  |  |  |  |  |  |  |  |  |  |  |  |  |  |  |
| England | 5544 | 99908 | 5.96 [5.78-6.14] | 4995 | 105829 | 4.83 [4.67-5.00] | 7037 | 159367 | 4.38 [4.25-4.51] | 8431 | 161537 | 5.56 [5.43-5.71] | -18.96 [-21.81, -16.11] | -9.11 [-12.01, -6.21] | 26.94 [23.97, 29.91] |
| **Sex**** |  |  |  |  |  |  |  |  |  |  |  |  |  |  |  |
| Male | 2405 | 43825 | 6.17 [5.91-6.44] | 2117 | 46269 | 4.87 [4.64-5.11] | 3029 | 69421 | 4.37 [4.19-4.56] | 3598 | 70727 | 5.51 [5.31-5.71] | -21.07 [-24.96, -17.18] | -10.27 [-14.58, -5.95] | 26.09 [21.74, 30.43] |
| Female | 3139 | 56083 | 5.75 [5.50-6.01] | 2878 | 59560 | 4.79 [4.57-5.03] | 4008 | 89944 | 4.39 [4.21-4.57] | 4833 | 90810 | 5.62 [5.42-5.82] | -16.70 [-20.87, -12.52] | -8.56 [-12.73, -4.38] | 27.79 [23.69, 31.89] |
| **Age**** |  |  |  |  |  |  |  |  |  |  |  |  |  |  |  |
| 18-24 | 463 | 6499 | 7.86 [7.26-8.50] | 411 | 6493 | 7.31 [6.75-7.90] | 574 | 8763 | 6.70 [6.25-7.17] | 851 | 9539 | 9.85 [9.33-10.38] | -7.12 [-14.50, 0.25] | -8.34 [-15.32, -1.37] | 46.87 [39.70, 54.03] |
| 25-34 | 930 | 13366 | 7.83 [7.35-8.32] | 775 | 13573 | 5.88 [5.47-6.32] | 1036 | 20212 | 5.15 [4.83-5.49] | 1287 | 21049 | 6.60 [6.25-6.96] | -24.90 [-30.52, -19.28] | -12.24 [-18.54, -5.95] | 27.96 [21.36, 34.56] |
| 35-44 | 964 | 17052 | 6.09 [5.65-6.56] | 837 | 17130 | 5.12 [4.71-5.55] | 1202 | 26687 | 4.60 [4.28-4.94] | 1365 | 26884 | 5.28 [4.94-5.63] | -16.09 [-23.15, -9.03] | -10.16 [-17.38, -2.93] | 14.78 [7.61, 21.96] |
| 45-54 | 1255 | 20634 | 6.41 [5.98-6.87] | 1100 | 21487 | 5.48 [5.08-5.90] | 1559 | 32403 | 4.96 [4.65-5.29] | 1812 | 32799 | 5.48 [5.16-5.82] | -14.51 [-21.06, -7.96] | -9.49 [-16.06, -2.92] | 10.48 [4.03, 16.94] |
| 55-64 | 1131 | 20404 | 5.92 [5.46-6.40] | 1074 | 21840 | 4.70 [4.30-5.14] | 1537 | 32870 | 4.33 [4.01-4.67] | 1767 | 33296 | 5.53 [5.17-5.89] | -20.61 [-27.87, -13.34] | -7.66 [-15.53, 0.21] | 27.48 [19.63, 35.33] |
| 65-74 | 568 | 15543 | 3.16 [2.76-3.59] | 594 | 17617 | 2.74 [2.37-3.14] | 787 | 26542 | 2.25 [1.96-2.56] | 981 | 26465 | 3.35 [3.03-3.70] | -13.61 [-25.95, -1.27] | -17.52 [-29.93, -5.11] | 48.89 [35.11, 62.67] |
| 75+ | 233 | 6410 | 3.31 [2.86-3.79] | 204 | 7689 | 1.61 [1.26-2.00] | 342 | 11890 | 2.01 [1.70-2.34] | 368 | 11505 | 2.63 [2.30-2.98] | -51.06 [-63.44, -38.67] | 24.84 [3.73, 45.96] | 30.85 [14.93, 46.77] |
| **Ethnicity**** |  |  |  |  |  |  |  |  |  |  |  |  |  |  |  |
| White | 4827 | 92737 | 5.01 [4.83-5.19] | 4384 | 98003 | 4.05 [3.89-4.22] | 6176 | 148227 | 3.63 [3.50-3.76] | 7450 | 150527 | 4.80 [4.66-4.94] | -19.16 [-22.55, -15.77] | -10.37 [-14.07, -6.67] | 32.23 [28.65, 35.81] |
| Mixed | 106 | 1347 | 8.92 [7.09-11.08] | 76 | 1308 | 6.19 [4.66-8.05] | 108 | 1865 | 5.69 [4.46-7.16] | 140 | 1959 | 7.18 [5.83-8.75] | -30.61 [-50.78, -10.43] | -7.92 [-31.99, 16.16] | 26.01 [1.93, 50.09] |
| Asian | 369 | 3658 | 11.86 [10.99-12.77] | 340 | 3930 | 11.23 [10.40-12.10] | 439 | 5518 | 9.70 [9.06-10.38] | 527 | 5425 | 11.65 [10.96-12.38] | -5.31 [-12.48, 1.85] | -13.62 [-20.21, -7.03] | 20.10 [13.20, 27.01] |
| Black | 135 | 900 | 17.34 [15.75-19.05] | 102 | 936 | 11.92 [10.56-13.42] | 154 | 1304 | 13.77 [12.56-15.06] | 119 | 1180 | 11.80 [10.66-13.04] | -31.26 [-39.91, -22.61] | 15.52 [4.53, 26.51] | -14.23 [-22.88, -5.59] |
| Other | 79 | 762 | 12.28 [10.21-14.66] | 70 | 939 | 8.25 [6.50-10.33] | 98 | 1393 | 8.25 [6.77-9.95] | 132 | 1315 | 13.36 [11.54-15.40] | -32.82 [-49.27, -16.37] | -0.12 [-20.85, 20.61] | 62.06 [41.21, 82.91] |
| **Region**** |  |  |  |  |  |  |  |  |  |  |  |  |  |  |  |
| North East | 196 | 3574 | 5.03 [4.30-5.85] | 202 | 4027 | 4.34 [3.66-5.10] | 296 | 6327 | 3.87 [3.33-4.46] | 359 | 6024 | 5.55 [4.95-6.21] | -13.92 [-28.43, 0.60] | -10.83 [-25.35, 3.69] | 43.41 [28.42, 58.40] |
| North West | 714 | 11996 | 6.65 [6.14-7.19] | 657 | 12995 | 5.25 [4.80-5.74] | 910 | 18616 | 4.50 [4.15-4.87] | 1304 | 19142 | 7.67 [7.24-8.12] | -21.05 [-28.42, -13.68] | -14.10 [-21.90, -6.29] | 70.22 [61.56, 78.89] |
| Yorkshire and The Humber | 284 | 6519 | 3.95 [3.46-4.48] | 306 | 7391 | 3.97 [3.50-4.49] | 399 | 10594 | 3.37 [3.01-3.77] | 588 | 11078 | 6.33 [5.88-6.81] | 0.76 [-11.65, 13.16] | -15.37 [-26.20, -4.53] | 88.13 [75.67, 100.59] |
| East Midlands | 601 | 12684 | 4.23 [3.71-4.80] | 533 | 13685 | 3.37 [2.90-3.89] | 756 | 20469 | 3.11 [2.73-3.52] | 942 | 20747 | 4.16 [3.75-4.60] | -20.33 [-32.39, -8.27] | -7.72 [-20.77, 5.34] | 33.76 [20.90, 46.62] |
| West Midlands | 547 | 9620 | 5.82 [5.28-6.40] | 592 | 10062 | 6.97 [6.41-7.57] | 672 | 15046 | 4.77 [4.37-5.19] | 879 | 15261 | 6.02 [5.59-6.48] | 19.93 [10.31, 29.55] | -31.71 [-38.74, -24.68] | 26.21 [17.40, 35.01] |
| East of England | 805 | 14433 | 5.09 [4.59-5.63] | 689 | 15189 | 4.02 [3.58-4.50] | 993 | 23174 | 3.69 [3.34-4.07] | 1061 | 23004 | 4.20 [3.83-4.59] | -20.83 [-30.26, -11.39] | -8.21 [-18.41, 1.99] | 13.55 [3.79, 23.31] |
| London | 1045 | 9547 | 12.96 [12.34-13.59] | 855 | 9872 | 9.38 [8.86-9.93] | 1265 | 15227 | 9.46 [9.03-9.91] | 1384 | 15544 | 9.54 [9.10-9.98] | -27.55 [-31.94, -23.15] | 0.75 [-4.37, 5.86] | 0.74 [-3.81, 5.29] |
| South East | 995 | 21979 | 3.92 [3.54-4.32] | 891 | 22632 | 3.09 [2.75-3.45] | 1325 | 34738 | 3.01 [2.74-3.30] | 1407 | 35170 | 3.25 [2.97-3.54] | -21.43 [-30.61, -12.24] | -2.27 [-12.30, 7.77] | 7.97 [-1.33, 17.28] |
| South West | 357 | 9556 | 2.79 [2.37-3.25] | 270 | 9976 | 1.28 [0.95-1.65] | 421 | 15176 | 1.62 [1.34-1.94] | 507 | 15567 | 2.29 [1.97-2.63] | -54.48 [-68.46, -40.50] | 27.34 [2.34, 52.34] | 40.74 [21.60, 59.88] |
| **IMD quintile** |  |  |  |  |  |  |  |  |  |  |  |  |  |  |  |
| Most deprived**: 1 | 682 | 10082 | 7.28 [6.84-7.74] | 639 | 10997 | 6.27 [5.86-6.69] | 827 | 15681 | 5.39 [5.08-5.71] | 999 | 15727 | 7.22 [6.87-7.57] | -13.87 [-19.64, -8.10] | -14.04 [-19.78, -8.29] | 33.95 [27.83, 40.07] |
| 2 | 947 | 16015 | 6.43 [6.03-6.85] | 855 | 16973 | 5.35 [4.98-5.73] | 1183 | 25206 | 4.82 [4.53-5.12] | 1332 | 25492 | 5.62 [5.32-5.94] | -16.80 [-22.71, -10.89] | -9.91 [-16.07, -3.74] | 16.80 [10.79, 22.82] |
| 3 | 1196 | 21474 | 5.87 [5.48-6.29] | 1027 | 23231 | 4.46 [4.12-4.82] | 1518 | 34548 | 4.37 [4.09-4.66] | 1815 | 34784 | 5.49 [5.18-5.80] | -24.19 [-30.49, -17.89] | -2.02 [-8.97, 4.93] | 25.40 [18.76, 32.04] |
| 4 | 1287 | 24840 | 5.22 [4.84-5.62] | 1182 | 25979 | 4.25 [3.91-4.61] | 1678 | 39595 | 3.84 [3.57-4.12] | 2007 | 40259 | 4.88 [4.59-5.19] | -18.58 [-25.48, -11.69] | -9.65 [-16.94, -2.35] | 27.08 [19.79, 34.38] |
| Least deprived: 5 | 1432 | 27497 | 4.99 [4.61-5.39] | 1292 | 28649 | 3.85 [3.52-4.21] | 1831 | 44337 | 3.51 [3.24-3.78] | 2278 | 45275 | 4.66 [4.37-4.97] | -22.65 [-29.86, -15.43] | -9.09 [-16.88, -1.30] | 32.76 [24.79, 40.74] |
| **Highest educational level reached** |  |  |  |  |  |  |  |  |  |  |  |  |  |  |  |
| No qualification | 420 | 9101 | 3.87 [3.38-4.42] | 398 | 10207 | 3.01 [2.58-3.48] | 557 | 14701 | 2.88 [2.52-3.26] | 643 | 13923 | 3.88 [3.47-4.31] | -22.22 [-34.37, -10.08] | -4.32 [-17.61, 8.97] | 34.72 [21.18, 48.26] |
| Other | 382 | 6758 | 5.12 [4.49-5.82] | 328 | 6979 | 3.98 [3.41-4.60] | 420 | 10552 | 3.11 [2.68-3.58] | 495 | 10309 | 4.10 [3.62-4.62] | -22.27 [-34.38, -10.16] | -21.86 [-34.92, -8.79] | 31.83 [17.04, 46.62] |
| GCSE | 1135 | 22204 | 4.47 [4.13-4.83] | 1024 | 23660 | 3.53 [3.22-3.85] | 1346 | 33802 | 3.11 [2.87-3.37] | 1632 | 34147 | 4.07 [3.80-4.35] | -21.03 [-28.19, -13.87] | -11.90 [-19.83, -3.97] | 30.87 [22.51, 39.23] |
| Post-GCSE qualification | 1515 | 27597 | 4.93 [4.61-5.26] | 1410 | 29290 | 4.11 [3.82-4.42] | 1985 | 43693 | 3.79 [3.56-4.03] | 2540 | 44526 | 5.19 [4.93-5.45] | -16.63 [-22.72, -10.55] | -8.03 [-14.36, -1.70] | 36.94 [30.61, 43.27] |
| Degree or above | 2062 | 33751 | 5.67 [5.37-5.99] | 1805 | 34911 | 4.54 [4.27-4.83] | 2677 | 55453 | 4.13 [3.92-4.35] | 3080 | 57487 | 4.77 [4.55-4.99] | -19.93 [-25.04, -14.81] | -9.03 [-14.32, -3.74] | 15.50 [10.41, 20.58] |
| **Gross household income** |  |  |  |  |  |  |  |  |  |  |  |  |  |  |  |
| £0-14,999 | 297 | 6482 | 3.83 [3.25-4.48] | 263 | 6936 | 2.88 [2.37-3.45] | 348 | 9978 | 2.52 [2.10-2.97] | 459 | 10363 | 3.65 [3.19-4.15] | -24.80 [-39.43, -10.18] | -12.50 [-29.17, 4.17] | 44.84 [27.38, 62.30] |
| £15,000-49,999 | 1650 | 31146 | 4.70 [4.40-5.00] | 1357 | 31362 | 3.53 [3.26-3.80] | 1890 | 45898 | 3.27 [3.06-3.50] | 2316 | 46899 | 4.26 [4.03-4.50] | -24.89 [-30.85, -18.94] | -7.08 [-13.88, -0.28] | 30.28 [23.55, 37.00] |
| £50,000-149,999 | 1707 | 27252 | 5.86 [5.52-6.21] | 1454 | 26945 | 4.81 [4.50-5.15] | 2047 | 39993 | 4.48 [4.22-4.75] | 2381 | 41196 | 5.28 [5.01-5.55] | -17.75 [-23.38, -12.12] | -7.07 [-13.10, -1.04] | 17.86 [12.05, 23.66] |
| >£150,000 | 284 | 3216 | 8.95 [7.83-10.19] | 193 | 3112 | 5.79 [4.83-6.87] | 328 | 5032 | 6.17 [5.38-7.03] | 377 | 5272 | 6.93 [6.13-7.81] | -35.42 [-47.49, -23.35] | 6.74 [-8.81, 22.28] | 12.32 [-0.81, 25.45] |
| **Employment** |  |  |  |  |  |  |  |  |  |  |  |  |  |  |  |
| Healthcare (patient-facing) | 379 | 3402 | 11.74 [10.51-13.06] | 389 | 3511 | 11.66 [10.46-12.96] | 578 | 5416 | 11.17 [10.21-12.20] | 589 | 6039 | 10.06 [9.19-11.00] | -0.60 [-11.07, 9.88] | -4.20 [-13.64, 5.23] | -9.94 [-18.17, -1.70] |
| Healthcare (other) | 73 | 1151 | 5.95 [4.43-7.83] | 62 | 1112 | 5.03 [3.58-6.85] | 113 | 1692 | 6.36 [5.04-7.91] | 133 | 1957 | 6.50 [5.26-7.95] | -15.29 [-42.52, 11.93] | 26.24 [-3.38, 55.86] | 2.36 [-18.87, 23.58] |
| Care home (client-facing) | 115 | 761 | 16.52 [13.67-19.80] | 83 | 727 | 12.07 [9.52-15.10] | 108 | 979 | 11.60 [9.42-14.15] | 94 | 853 | 11.59 [9.27-14.34] | -26.94 [-44.19, -9.69] | -3.89 [-24.77, 16.98] | -0.09 [-20.69, 20.52] |
| Care home (other) | 12 | 146 | 8.22 [4.05-14.96] | 12 | 224 | 4.77 [2.04-9.31] | 23 | 257 | 9.10 [5.59-14.06] | 21 | 233 | 9.17 [5.51-14.44] | -41.97 [-95.86, 11.92] | 90.78 [11.32, 170.23] | 0.77 [-45.49, 47.03] |
| Other essential worker | 1209 | 19927 | 5.62 [5.23-6.03] | 1019 | 19615 | 4.57 [4.21-4.96] | 1463 | 29572 | 4.27 [3.98-4.58] | 1799 | 31787 | 5.13 [4.83-5.44] | -18.68 [-25.44, -11.92] | -6.56 [-13.79, 0.66] | 19.91 [13.11, 26.70] |
| Other worker | 2189 | 37855 | 5.28 [5.00-5.57] | 1982 | 40782 | 4.17 [3.92-4.43] | 2704 | 60731 | 3.68 [3.48-3.88] | 3197 | 59632 | 4.77 [4.56-4.99] | -21.02 [-25.95, -16.10] | -11.99 [-17.27, -6.71] | 29.89 [24.46, 35.33] |
| Not in employment | 1516 | 35737 | 3.42 [3.18-3.68] | 1412 | 39030 | 2.67 [2.45-2.90] | 1988 | 59369 | 2.35 [2.18-2.53] | 2500 | 59546 | 3.37 [3.18-3.57] | -21.93 [-28.65, -15.20] | -11.99 [-19.48, -4.49] | 43.40 [35.74, 51.06] |
| **Employment (detailed)** |  |  |  |  |  |  |  |  |  |  |  |  |  |  |  |
| Full time | 2744 | 41780 | 6.23 [5.94-6.52] | 2425 | 42816 | 5.14 [4.88-5.41] | 3491 | 64672 | 4.82 [4.61-5.03] | 4055 | 66389 | 5.67 [5.46-5.89] | -17.66 [-21.99, -13.32] | -6.03 [-10.51, -1.56] | 17.63 [13.28, 21.99] |
| Part time | 714 | 12510 | 5.19 [4.72-5.70] | 663 | 13455 | 4.25 [3.82-4.71] | 898 | 19599 | 3.83 [3.49-4.20] | 1065 | 19612 | 4.86 [4.48-5.25] | -18.11 [-26.78, -9.44] | -9.88 [-19.06, -0.71] | 26.89 [17.49, 36.29] |
| Self-employed | 570 | 9859 | 5.28 [4.74-5.86] | 495 | 10525 | 3.98 [3.51-4.49] | 660 | 15726 | 3.37 [3.01-3.76] | 811 | 15990 | 4.42 [4.03-4.85] | -24.62 [-34.28, -14.96] | -15.33 [-25.88, -4.77] | 31.16 [19.88, 42.43] |
| Govt supported training | 4 | 36 | 11.70 [3.62-28.81] | 1 | 41 | 1.25 [0.00-13.49] | 2 | 45 | 3.67 [0.00-16.18] | 6 | 61 | 10.16 [3.84-22.22] | -89.32 [-169.49, -9.15] | 192.80 [-315.20, 700.80] | 177.38 [-39.78, 394.55] |
| Unemployed | 110 | 1975 | 5.02 [3.91-6.35] | 113 | 2326 | 4.17 [3.20-5.31] | 144 | 3430 | 3.37 [2.62-4.24] | 187 | 3480 | 4.79 [3.94-5.75] | -16.93 [-39.04, 5.18] | -19.18 [-41.01, 2.64] | 41.84 [17.21, 66.47] |
| Retired | 751 | 21730 | 2.48 [2.19-2.78] | 743 | 24568 | 1.96 [1.71-2.22] | 1062 | 37468 | 1.73 [1.53-1.94] | 1269 | 37125 | 2.43 [2.21-2.66] | -21.37 [-32.26, -10.48] | -11.73 [-23.47, 0.00] | 41.04 [28.90, 53.18] |
| Student | 152 | 2086 | 7.09 [5.84-8.53] | 108 | 1565 | 6.63 [5.24-8.27] | 152 | 2419 | 5.88 [4.80-7.13] | 337 | 2950 | 12.08 [10.75-13.52] | -6.63 [-26.23, 12.98] | -11.31 [-31.22, 8.60] | 105.27 [84.01, 126.53] |
| Looking after home/family | 204 | 4245 | 4.10 [3.38-4.93] | 163 | 3898 | 3.35 [2.65-4.16] | 235 | 6074 | 2.97 [2.42-3.59] | 246 | 5949 | 3.30 [2.72-3.94] | -18.54 [-36.83, -0.24] | -11.04 [-30.75, 8.66] | 10.77 [-8.75, 30.30] |
| Sick/disabled | 69 | 1766 | 3.02 [2.05-4.24] | 73 | 1943 | 2.84 [1.93-3.97] | 89 | 2831 | 2.10 [1.40-2.96] | 102 | 2848 | 2.63 [1.88-3.53] | -5.96 [-40.07, 28.15] | -26.06 [-57.04, 4.93] | 25.24 [-11.90, 62.38] |
| Other | 91 | 1837 | 4.28 [3.20-5.59] | 83 | 1883 | 3.62 [2.61-4.86] | 104 | 2541 | 3.24 [2.40-4.26] | 131 | 2672 | 4.22 [3.31-5.29] | -15.19 [-41.59, 11.21] | -10.77 [-38.40, 16.85] | 30.25 [1.54, 58.95] |
| Prefer not to say | 135 | 2062 | 6.20 [5.01-7.59] | 128 | 2806 | 3.81 [2.95-4.82] | 200 | 4561 | 3.60 [2.93-4.36] | 222 | 4461 | 4.31 [3.59-5.13] | -38.55 [-56.13, -20.97] | -5.51 [-26.77, 15.75] | 19.72 [-0.28, 39.72] |
| **Employment (sector)** |  |  |  |  |  |  |  |  |  |  |  |  |  |  |  |
| Home delivery | 8 | 102 | 7.76 [3.17-16.05] | 36 | 1026 | 2.54 [1.38-4.12] | 76 | 1784 | 3.45 [2.43-4.70] | 102 | 1805 | 5.12 [3.95-6.52] | -67.27 [-124.36, -10.18] | 35.83 [-12.20, 83.86] | 48.41 [14.20, 82.61] |
| Retail | 30 | 513 | 5.36 [3.29-8.22] | 284 | 5810 | 4.20 [3.57-4.91] | 258 | 6280 | 3.26 [2.70-3.89] | 343 | 6406 | 4.76 [4.13-5.46] | -21.64 [-54.29, 11.01] | -22.14 [-36.67, -7.62] | 45.71 [26.99, 64.42] |
| Emergency services, prisons, coastguard | 3 | 43 | 6.72 [1.21-20.73] | 65 | 934 | 6.70 [4.94-8.88] | 69 | 1343 | 4.50 [3.23-6.09] | 100 | 1481 | 6.45 [5.04-8.13] | -0.30 [-96.43, 95.83] | -32.69 [-57.61, -7.76] | 43.11 [11.11, 75.11] |
| Public transport or taxi | 4 | 33 | 12.92 [4.12-31.24] | 45 | 757 | 5.48 [3.70-7.79] | 69 | 1177 | 5.38 [3.93-7.17] | 84 | 1161 | 7.03 [5.40-9.00] | -57.66 [-130.03, 14.71] | -1.82 [-34.49, 30.84] | 31.04 [0.19, 61.90] |
| Teacher or childcare | 30 | 346 | 8.76 [5.71-12.90] | 365 | 5987 | 5.66 [4.96-6.42] | 566 | 11465 | 4.26 [3.80-4.76] | 704 | 11448 | 5.72 [5.21-6.27] | -35.39 [-64.04, -6.74] | -24.56 [-35.16, -13.96] | 34.04 [22.54, 45.54] |
| Armed forces | 0 | 9 | 0.00 [0.00-26.16] | 3 | 97 | 2.04 [0.00-8.80] | 10 | 132 | 7.44 [3.33-14.44] | 4 | 143 | 1.68 [0.00-6.71] | Inf [Inf, Inf] | 265.20 [34.31, 496.08] | -77.42 [-136.16, -18.68] |
| Other | 101 | 1597 | 5.93 [4.62-7.50] | 1029 | 21495 | 4.08 [3.75-4.43] | 1962 | 40297 | 4.18 [3.93-4.44] | 2243 | 39062 | 5.23 [4.96-5.51] | -31.20 [-48.40, -14.00] | 2.45 [-4.66, 9.56] | 25.12 [18.90, 31.34] |
| Not working outside home | 1293 | 32699 | 3.08 [2.83-3.34] | 1308 | 26118 | 4.35 [4.04-4.67] | 1380 | 30722 | 3.73 [3.45-4.01] | 1673 | 33005 | 4.42 [4.14-4.71] | 41.56 [32.47, 50.65] | -14.48 [-21.15, -7.82] | 18.77 [11.53, 26.01] |
| **Household size** |  |  |  |  |  |  |  |  |  |  |  |  |  |  |  |
| 1 | 720 | 15052 | 4.08 [3.68-4.50] | 671 | 16777 | 3.13 [2.79-3.50] | 953 | 24735 | 2.96 [2.67-3.25] | 1141 | 24889 | 3.84 [3.53-4.16] | -23.04 [-32.11, -13.97] | -5.75 [-15.97, 4.47] | 29.73 [19.93, 39.53] |
| 2 | 1784 | 36413 | 4.22 [3.95-4.49] | 1593 | 39252 | 3.20 [2.97-3.44] | 2273 | 59922 | 2.88 [2.70-3.07] | 2822 | 62314 | 3.77 [3.58-3.97] | -24.17 [-30.09, -18.25] | -10.00 [-16.56, -3.44] | 30.90 [24.31, 37.50] |
| 3 | 1158 | 19734 | 5.38 [5.00-5.79] | 1065 | 20898 | 4.45 [4.10-4.82] | 1484 | 31429 | 4.00 [3.73-4.29] | 1794 | 32057 | 5.06 [4.76-5.37] | -17.29 [-24.16, -10.41] | -10.34 [-17.30, -3.37] | 26.50 [19.25, 33.75] |
| 4 | 1204 | 19611 | 5.71 [5.32-6.13] | 1077 | 20110 | 4.77 [4.40-5.15] | 1549 | 30208 | 4.49 [4.20-4.80] | 1724 | 29491 | 5.36 [5.04-5.69] | -16.46 [-23.12, -9.81] | -5.87 [-12.79, 1.05] | 19.38 [12.69, 26.06] |
| 5 | 447 | 6403 | 6.72 [6.00-7.51] | 392 | 6174 | 5.96 [5.26-6.73] | 529 | 9354 | 5.13 [4.58-5.71] | 595 | 8938 | 6.33 [5.73-6.98] | -11.31 [-22.02, -0.60] | -13.93 [-24.66, -3.19] | 23.59 [12.28, 34.89] |
| 6 | 152 | 1848 | 8.22 [6.82-9.84] | 124 | 1822 | 6.51 [5.23-8.02] | 162 | 2575 | 5.89 [4.84-7.10] | 209 | 2537 | 8.24 [7.02-9.60] | -20.80 [-38.08, -3.53] | -9.68 [-28.73, 9.37] | 39.90 [19.86, 59.93] |
| 7+ | 79 | 827 | 9.82 [7.63-12.47] | 73 | 796 | 9.36 [7.18-12.02] | 87 | 1130 | 7.59 [5.88-9.64] | 146 | 1311 | 11.73 [9.81-13.92] | -4.68 [-28.62, 19.25] | -18.91 [-41.45, 3.63] | 54.55 [29.38, 79.71] |
| **One or more children (under 18) in household** |  |  |  |  |  |  |  |  |  |  |  |  |  |  |  |
| No | 3647 | 67515 | 4.82 [4.62-5.03] | 3338 | 73258 | 3.80 [3.62-3.99] | 4693 | 109481 | 3.48 [3.34-3.62] | 5699 | 111275 | 4.48 [4.33-4.64] | -20.95 [-24.90, -17.01] | -8.68 [-12.89, -4.47] | 28.74 [24.43, 33.05] |
| Yes | 1897 | 32373 | 5.37 [5.07-5.69] | 1657 | 32571 | 4.44 [4.16-4.74] | 2343 | 49867 | 3.97 [3.75-4.20] | 2732 | 50262 | 4.86 [4.63-5.10] | -17.13 [-22.53, -11.73] | -10.59 [-16.22, -4.95] | 22.42 [16.62, 28.21] |
| **History of COVID-19 symptoms** |  |  |  |  |  |  |  |  |  |  |  |  |  |  |  |
| No symptoms | 87 | 683 | 13.66 [10.90-16.93] | 67 | 695 | 9.93 [7.55-12.85] | 107 | 1112 | 9.91 [7.98-12.17] | 188 | 1247 | 16.48 [14.21-19.00] | -27.31 [-47.51, -7.10] | -0.30 [-23.67, 23.06] | 66.40 [44.30, 88.50] |
| Mild symptoms | 1025 | 6071 | 18.65 [17.54-19.82] | 821 | 5396 | 16.64 [15.52-17.83] | 1189 | 8481 | 15.20 [14.33-16.11] | 1422 | 8506 | 18.45 [17.52-19.43] | -10.78 [-16.78, -4.77] | -8.65 [-14.66, -2.64] | 21.45 [15.53, 27.37] |
| Moderate symptoms | 1850 | 9261 | 22.38 [21.42-23.38] | 1670 | 8866 | 21.01 [20.04-22.00] | 2326 | 14298 | 17.91 [17.20-18.65] | 2720 | 14019 | 21.69 [20.91-22.49] | -6.17 [-10.46, -1.88] | -14.71 [-18.71, -10.71] | 21.05 [16.92, 25.18] |
| Severe symtoms | 884 | 3501 | 28.73 [27.03-30.50] | 872 | 3654 | 27.07 [25.43-28.76] | 1271 | 6348 | 22.44 [21.27-23.64] | 1328 | 5841 | 25.71 [24.43-27.02] | -5.81 [-11.59, -0.03] | -17.10 [-22.31, -11.90] | 14.62 [9.22, 20.01] |
| **BMI** |  |  |  |  |  |  |  |  |  |  |  |  |  |  |  |
| Underweight (<18.5) | 65 | 1236 | 4.65 [3.31-6.32] | 46 | 1416 | 2.23 [1.26-3.50] | 62 | 1993 | 2.06 [1.25-3.09] | 89 | 1975 | 3.74 [2.74-4.96] | -52.04 [-79.78, -24.30] | -7.62 [-52.02, 36.77] | 82.04 [33.98, 130.10] |
| Normal (18.5-24.9) | 1919 | 36191 | 4.70 [4.43-4.99] | 1722 | 38348 | 3.72 [3.48-3.98] | 2394 | 57658 | 3.32 [3.12-3.52] | 2965 | 59112 | 4.36 [4.15-4.57] | -20.85 [-26.38, -15.32] | -11.02 [-16.94, -5.11] | 31.63 [25.60, 37.65] |
| Overweight (25-29.9) | 1826 | 31818 | 5.23 [4.93-5.54] | 1605 | 33467 | 4.09 [3.82-4.37] | 2285 | 50215 | 3.80 [3.58-4.02] | 2664 | 51230 | 4.58 [4.35-4.81] | -21.61 [-26.96, -16.25] | -7.33 [-13.20, -1.47] | 20.53 [14.74, 26.32] |
| Obese (>=30) | 1161 | 19855 | 5.36 [4.98-5.76] | 1121 | 20908 | 4.77 [4.41-5.15] | 1545 | 30244 | 4.47 [4.18-4.77] | 1816 | 30311 | 5.53 [5.22-5.86] | -11.01 [-17.91, -4.10] | -6.29 [-13.21, 0.63] | 23.71 [17.00, 30.43] |
| **Current smoker** |  |  |  |  |  |  |  |  |  |  |  |  |  |  |  |
| Yes | 433 | 10635 | 3.22 [2.79-3.69] | 375 | 11501 | 2.24 [1.87-2.65] | 475 | 16389 | 1.81 [1.51-2.13] | 580 | 15884 | 2.71 [2.37-3.08] | -30.43 [-43.17, -17.70] | -19.20 [-34.38, -4.02] | 49.72 [32.04, 67.40] |
| No | 5059 | 88290 | 5.22 [5.03-5.40] | 4580 | 93580 | 4.21 [4.05-4.38] | 6513 | 141756 | 3.85 [3.72-3.98] | 7777 | 144378 | 4.80 [4.66-4.94] | -19.54 [-22.80, -16.28] | -8.55 [-12.11, -4.99] | 25.19 [21.82, 28.57] |
| **Population density quintile** |  |  |  |  |  |  |  |  |  |  |  |  |  |  |  |
| 1 | 806 | 19719 | 3.24 [2.92-3.58] | 677 | 20186 | 2.35 [2.07-2.66] | 962 | 30556 | 2.11 [1.88-2.35] | 1161 | 31036 | 2.82 [2.57-3.08] | -27.47 [-37.04, -17.90] | -10.21 [-21.28, 0.85] | 33.65 [22.27, 45.02] |
| 2 | 996 | 19459 | 4.48 [4.12-4.86] | 889 | 20021 | 3.66 [3.33-4.02] | 1269 | 30692 | 3.29 [3.03-3.57] | 1536 | 30893 | 4.30 [4.02-4.60] | -18.30 [-26.12, -10.49] | -10.11 [-18.31, -1.91] | 30.70 [22.49, 38.91] |
| 3 | 1023 | 19746 | 4.56 [4.19-4.94] | 931 | 20025 | 3.91 [3.57-4.28] | 1252 | 30267 | 3.30 [3.03-3.57] | 1531 | 30787 | 4.30 [4.02-4.60] | -13.82 [-21.49, -6.14] | -15.86 [-23.79, -7.93] | 30.30 [22.12, 38.48] |
| 4 | 1144 | 20171 | 5.15 [4.77-5.54] | 954 | 20181 | 4.01 [3.67-4.37] | 1410 | 30066 | 3.96 [3.68-4.26] | 1560 | 30304 | 4.52 [4.22-4.82] | -21.94 [-28.93, -14.95] | -1.25 [-8.98, 6.48] | 14.14 [6.82, 21.46] |
| 5 | 1575 | 20813 | 7.43 [7.01-7.87] | 1544 | 25416 | 5.63 [5.29-5.99] | 2144 | 37786 | 5.15 [4.87-5.44] | 2643 | 38517 | 6.58 [6.28-6.89] | -24.23 [-29.48, -18.98] | -8.70 [-14.21, -3.20] | 27.96 [22.33, 33.59] |
| **History of COVID-19** |  |  |  |  |  |  |  |  |  |  |  |  |  |  |  |
| Positive test | 277 | 341 | 96.18 [90.78-100.00] | 274 | 360 | 90.01 [84.39-94.97] | 474 | 750 | 74.46 [70.23-78.53] | 1094 | 1765 | 72.99 [70.23-75.69] | -6.42 [-11.63, -1.20] | -17.28 [-22.43, -12.12] | -1.97 [-6.58, 2.63] |
| Suspected by doctor | 353 | 1144 | 35.49 [32.35-38.79] | 348 | 1214 | 32.85 [29.87-35.99] | 470 | 1811 | 29.58 [27.21-32.07] | 482 | 1850 | 29.70 [27.36-32.17] | -7.41 [-16.03, 1.21] | -9.98 [-18.20, -1.77] | 0.41 [-7.57, 8.38] |
| Suspected by respondent | 3118 | 17893 | 19.31 [18.65-19.99] | 2742 | 16914 | 17.85 [17.19-18.52] | 3867 | 27546 | 15.23 [14.74-15.73] | 3950 | 25777 | 16.78 [16.25-17.31] | -7.61 [-10.98, -4.25] | -14.62 [-17.82, -11.43] | 10.11 [6.83, 13.39] |
| No | 1698 | 80390 | 0.86 [0.74-0.98] | 1565 | 87217 | 0.48 [0.37-0.58] | 2144 | 129126 | 0.31 [0.23-0.40] | 2773 | 131924 | 0.85 [0.75-0.94] | -45.35 [-58.14, -32.56] | -33.33 [-52.08, -14.58] | 170.97 [141.94, 200.00] |
| **Time since positive PCR test** |  |  |  |  |  |  |  |  |  |  |  |  |  |  |  |
| <30 days ago | 69 | 90 | 90.68 [78.97-99.76] | 15 | 28 | 62.86 [41.46-83.21] | 43 | 109 | 45.84 [35.40-57.15] | 431 | 798 | 63.39 [59.21-67.52] | -30.69 [-49.39, -11.99] | -27.08 [-54.31, 0.16] | 38.29 [20.53, 56.04] |
| 30-60 days ago | 74 | 95 | 92.16 [80.91-100.00] | 48 | 65 | 87.28 [73.07-98.29] | 42 | 61 | 81.27 [66.28-93.57] | 168 | 218 | 91.16 [83.91-97.29] | -5.28 [-17.49, 6.92] | -6.90 [-21.91, 8.11] | 12.17 [-1.00, 25.34] |
| 61-90 days ago | 106 | 125 | 100.00 [91.66-100.00] | 68 | 92 | 87.37 [75.56-96.86] | 49 | 95 | 60.46 [48.51-72.25] | 38 | 66 | 67.68 [53.21-81.15] | -13.12 [-22.22, -4.02] | -30.79 [-43.58, -17.99] | 11.96 [-9.44, 33.36] |
| 90+ days ago | 28 | 31 | 100.00 [88.80-100.00] | 143 | 175 | 96.76 [89.07-100.00] | 340 | 485 | 82.77 [77.69-87.48] | 457 | 683 | 78.93 [74.57-83.06] | -10.37 [-20.23, -0.51] | -14.46 [-20.48, -8.43] | -4.64 [-10.04, 0.76] |
| **Time since COVID-19 symptom onset** |  |  |  |  |  |  |  |  |  |  |  |  |  |  |  |
| <30 days ago | 181 | 1052 | 19.04 [16.44-21.93] | 97 | 756 | 13.77 [11.12-16.87] | 124 | 1280 | 9.98 [8.18-12.09] | 582 | 2247 | 29.52 [27.39-31.75] | -27.73 [-42.12, -13.34] | -27.45 [-44.81, -10.09] | 195.69 [175.45, 215.93] |
| 30-60 days ago | 350 | 1323 | 30.19 [27.41-33.13] | 70 | 355 | 22.07 [17.49-27.44] | 63 | 414 | 16.65 [12.87-21.20] | 138 | 725 | 21.25 [18.00-24.88] | -26.90 [-40.01, -13.78] | -24.56 [-44.86, -4.26] | 27.57 [5.23, 49.91] |
| 61-90 days ago | 1884 | 5085 | 42.95 [41.37-44.56] | 229 | 873 | 29.92 [26.53-33.55] | 73 | 451 | 17.81 [14.07-22.25] | 50 | 334 | 16.35 [12.22-21.44] | -30.34 [-36.53, -24.14] | -40.44 [-52.87, -28.01] | -8.25 [-32.12, 15.61] |
| 91-120 days ago | 783 | 4832 | 17.84 [16.62-19.12] | 1251 | 3297 | 44.03 [42.05-46.04] | 157 | 799 | 21.99 [18.85-25.48] | 56 | 314 | 19.80 [15.18-25.36] | 146.80 [137.72, 155.89] | -50.03 [-56.10, -43.97] | -9.96 [-29.01, 9.10] |
| 121-150 days ago | 120 | 2649 | 3.77 [2.89-4.81] | 1144 | 5289 | 24.37 [23.06-25.73] | 668 | 2131 | 36.08 [33.75-38.49] | 113 | 468 | 27.40 [23.00-32.32] | 546.42 [516.45, 576.39] | 48.05 [40.34, 55.77] | -24.03 [-34.04, -14.02] |
| >150 days ago | 104 | 2006 | 4.56 [3.49-5.84] | 227 | 5177 | 3.60 [2.96-4.31] | 3186 | 20174 | 17.34 [16.74-17.95] | 3801 | 20191 | 20.99 [20.35-21.65] | -21.05 [-41.45, -0.66] | 381.94 [364.72, 399.17] | 21.11 [17.59, 24.63] |
| **Contact with COVID-19 case** |  |  |  |  |  |  |  |  |  |  |  |  |  |  |  |
| Yes, with confirmed case | 742 | 3946 | 20.97 [19.54-22.47] | 753 | 4543 | 18.28 [17.01-19.62] | 1107 | 7793 | 15.43 [14.52-16.38] | 1890 | 12871 | 16.01 [15.28-16.75] | -12.78 [-19.22, -6.34] | -15.59 [-21.61, -9.57] | 3.69 [-1.62, 9.01] |
| Yes, with suspected case | 896 | 5307 | 18.65 [17.47-19.90] | 842 | 5115 | 18.15 [16.95-19.40] | 1121 | 7362 | 16.66 [15.69-17.67] | 1111 | 6920 | 17.66 [16.64-18.72] | -2.73 [-9.12, 3.65] | -8.15 [-14.10, -2.20] | 5.94 [0.00, 11.88] |
| No | 3906 | 90655 | 3.50 [3.35-3.67] | 3400 | 96171 | 2.57 [2.43-2.72] | 4809 | 144211 | 2.33 [2.22-2.44] | 5430 | 141746 | 2.93 [2.81-3.05] | -26.57 [-30.86, -22.29] | -9.73 [-14.40, -5.06] | 25.75 [21.03, 30.47] |
| **Number of existing health conditions** |  |  |  |  |  |  |  |  |  |  |  |  |  |  |  |
| >1 | 1097 | 22127 | 4.29 [3.95-4.64] | 1112 | 24429 | 3.80 [3.49-4.12] | 962 | 24548 | 3.03 [2.75-3.34] | 1216 | 25468 | 4.07 [3.76-4.39] | -11.42 [-18.88, -3.96] | -20.00 [-27.89, -12.11] | 33.66 [23.76, 43.56] |
| 1 | 1558 | 28308 | 4.94 [4.63-5.27] | 1407 | 30313 | 3.91 [3.63-4.20] | 1721 | 41156 | 3.35 [3.12-3.59] | 2022 | 41362 | 4.20 [3.96-4.46] | -21.05 [-27.13, -14.98] | -14.07 [-20.46, -7.67] | 25.37 [18.21, 32.54] |
| 0 | 2889 | 49473 | 5.35 [5.10-5.60] | 2476 | 51087 | 4.15 [3.93-4.38] | 4354 | 93663 | 3.91 [3.75-4.08] | 5193 | 94707 | 4.92 [4.75-5.10] | -22.24 [-26.54, -17.94] | -5.78 [-10.36, -1.20] | 25.58 [21.48, 29.67] |
| **Care home resident** |  |  |  |  |  |  |  |  |  |  |  |  |  |  |  |
| Yes | 6 | 131 | 3.83 [0.86-9.92] | 18 | 259 | 6.69 [3.66-11.23] | 25 | 348 | 6.97 [4.23-10.83] | 25 | 313 | 7.94 [4.90-12.20] | 74.67 [-27.94, 177.28] | 4.04 [-47.09, 55.16] | 14.06 [-34.15, 62.27] |
| No | 5538 | 99777 | 5.00 [4.83-5.17] | 4977 | 105570 | 3.99 [3.84-4.15] | 7012 | 159019 | 3.63 [3.51-3.75] | 8406 | 161224 | 4.60 [4.47-4.73] | -20.20 [-23.40, -17.00] | -9.02 [-12.53, -5.51] | 26.45 [23.14, 29.75] |
| **Reported symptoms of previous COVID-19 case** |  |  |  |  |  |  |  |  |  |  |  |  |  |  |  |
| No symptoms | 1785 | 81076 | 0.97 [0.85-1.09] | 1632 | 87913 | 0.55 [0.44-0.66] | 2262 | 130401 | 0.40 [0.32-0.49] | 2982 | 133318 | 1.01 [0.91-1.11] | -42.27 [-53.61, -30.93] | -27.27 [-43.64, -10.91] | 152.50 [130.00, 175.00] |
| Atypical symptoms only | 353 | 3500 | 10.46 [9.31-11.72] | 298 | 3243 | 9.38 [8.24-10.64] | 474 | 5145 | 9.41 [8.50-10.40] | 571 | 5011 | 12.04 [11.02-13.14] | -10.33 [-21.51, 0.86] | 0.21 [-10.98, 11.41] | 27.95 [17.53, 38.36] |
| Screening symptoms | 3406 | 15332 | 25.08 [24.29-25.88] | 3065 | 14673 | 23.48 [22.70-24.28] | 4301 | 23821 | 20.07 [19.48-20.66] | 4878 | 23208 | 23.64 [23.01-24.27] | -6.34 [-9.41, -3.27] | -14.52 [-17.42, -11.63] | 17.79 [14.85, 20.73] |
| **Number of children (under 18) in household** |  |  |  |  |  |  |  |  |  |  |  |  |  |  |  |
| 0 | 3647 | 67515 | 4.82 [4.62-5.03] | 3338 | 73258 | 3.80 [3.62-3.99] | 4693 | 109481 | 3.48 [3.34-3.62] | 5699 | 111275 | 4.48 [4.33-4.64] | -20.95 [-24.90, -17.01] | -8.68 [-12.89, -4.47] | 28.74 [24.43, 33.05] |
| 1 | 851 | 14471 | 5.40 [4.95-5.87] | 777 | 14639 | 4.71 [4.28-5.16] | 1074 | 22130 | 4.16 [3.83-4.51] | 1238 | 22282 | 5.01 [4.65-5.38] | -12.78 [-20.93, -4.63] | -11.68 [-19.75, -3.61] | 20.43 [12.26, 28.61] |
| 2 | 808 | 13803 | 5.37 [4.91-5.85] | 659 | 13955 | 4.00 [3.59-4.44] | 978 | 21471 | 3.80 [3.47-4.15] | 1127 | 21645 | 4.59 [4.24-4.95] | -25.33 [-33.52, -17.13] | -5.00 [-14.25, 4.25] | 20.79 [11.84, 29.74] |
| >2 | 238 | 4099 | 5.31 [4.50-6.22] | 221 | 3977 | 5.01 [4.20-5.92] | 291 | 6266 | 3.91 [3.31-4.57] | 367 | 6335 | 5.29 [4.63-6.02] | -5.65 [-21.47, 10.17] | -22.16 [-36.73, -7.58] | 35.55 [19.18, 51.92] |
| **COVID outcome severity** |  |  |  |  |  |  |  |  |  |  |  |  |  |  |  |
| No treatment | 2308 | 11749 | 21.98 [21.13-22.86] | 2003 | 11058 | 20.14 [19.29-21.01] | 2936 | 17898 | 18.08 [17.43-18.74] | 3488 | 17809 | 21.91 [21.22-22.62] | -8.42 [-12.24, -4.60] | -10.23 [-13.95, -6.50] | 21.29 [17.64, 24.94] |
| Sought medical care | 1201 | 5925 | 22.73 [21.53-23.99] | 1125 | 5754 | 21.87 [20.66-23.13] | 1475 | 9237 | 17.55 [16.67-18.47] | 1589 | 8636 | 20.48 [19.51-21.48] | -3.78 [-9.06, 1.50] | -19.75 [-24.55, -14.95] | 16.70 [11.45, 21.94] |
| Hospital admission | 54 | 189 | 32.74 [25.55-40.95] | 66 | 206 | 36.91 [29.69-44.93] | 95 | 344 | 31.59 [26.22-37.55] | 107 | 369 | 33.25 [27.96-39.07] | 12.80 [-10.17, 35.77] | -14.44 [-32.27, 3.39] | 5.25 [-12.12, 22.63] |
| ICU | 13 | 27 | 56.32 [35.35-77.85] | 16 | 35 | 53.39 [35.02-72.78] | 29 | 58 | 58.55 [43.54-73.57] | 20 | 49 | 47.49 [32.31-64.28] | -5.22 [-42.13, 31.69] | 9.66 [-22.87, 42.20] | -18.89 [-45.60, 7.82] |
| **Number of children under 5 in household** |  |  |  |  |  |  |  |  |  |  |  |  |  |  |  |
| 0 | 4986 | 90458 | 4.95 [4.78-5.14] | 4558 | 96260 | 4.02 [3.86-4.18] | 6381 | 145252 | 3.61 [3.48-3.73] | 7734 | 147006 | 4.65 [4.52-4.79] | -18.79 [-22.22, -15.35] | -10.45 [-13.93, -6.97] | 29.09 [25.48, 32.69] |
| 1 | 451 | 7906 | 5.19 [4.60-5.83] | 360 | 8038 | 3.71 [3.19-4.28] | 570 | 11965 | 4.05 [3.61-4.53] | 589 | 12143 | 4.16 [3.71-4.64] | -28.32 [-39.31, -17.34] | 9.16 [-4.04, 22.37] | 2.72 [-8.40, 13.83] |
| 2 | 105 | 1487 | 6.82 [5.38-8.53] | 72 | 1475 | 4.19 [3.00-5.67] | 84 | 2069 | 3.20 [2.28-4.34] | 104 | 2332 | 3.69 [2.76-4.79] | -38.56 [-59.24, -17.89] | -23.63 [-51.07, 3.82] | 15.31 [-15.62, 46.25] |
| >2 | 2 | 57 | 2.54 [0.00-12.68] | 5 | 56 | 9.07 [2.98-21.51] | 2 | 81 | 1.29 [0.00-8.63] | 4 | 56 | 6.92 [1.70-18.77] | 257.09 [-32.68, 546.85] | -85.78 [-160.75, -10.80] | 436.43 [-48.84, 921.71] |
| **Number of children aged 5 to 10 in household** |  |  |  |  |  |  |  |  |  |  |  |  |  |  |  |
| 0 | 4880 | 87764 | 5.01 [4.83-5.20] | 4441 | 93603 | 4.03 [3.87-4.20] | 6200 | 140455 | 3.63 [3.50-3.76] | 7469 | 142778 | 4.62 [4.48-4.76] | -19.76 [-23.15, -16.37] | -9.68 [-13.15, -6.20] | 27.00 [23.42, 30.58] |
| 1 | 494 | 9383 | 4.66 [4.13-5.22] | 416 | 9482 | 3.60 [3.12-4.12] | 652 | 14560 | 3.71 [3.32-4.13] | 738 | 14558 | 4.42 [4.01-4.86] | -22.53 [-33.48, -11.59] | 3.06 [-9.17, 15.28] | 19.14 [8.09, 30.19] |
| 2 | 164 | 2610 | 5.88 [4.84-7.08] | 129 | 2597 | 4.30 [3.37-5.39] | 171 | 4078 | 3.37 [2.67-4.16] | 205 | 3947 | 4.57 [3.79-5.46] | -26.87 [-44.56, -9.18] | -21.86 [-41.86, -1.86] | 35.91 [13.06, 58.75] |
| >2 | 6 | 151 | 3.10 [0.52-8.43] | 9 | 147 | 5.69 [2.23-11.84] | 14 | 274 | 4.47 [2.01-8.42] | 19 | 254 | 7.33 [4.15-12.03] | 83.55 [-49.68, 216.77] | -21.44 [-89.63, 46.75] | 63.98 [-13.20, 141.16] |
| **Number of children aged 11 to 17 in household** |  |  |  |  |  |  |  |  |  |  |  |  |  |  |  |
| 0 | 4624 | 84694 | 4.89 [4.71-5.08] | 4164 | 90623 | 3.85 [3.69-4.02] | 5869 | 135750 | 3.52 [3.39-3.65] | 7052 | 138160 | 4.46 [4.32-4.60] | -21.47 [-24.95, -18.00] | -8.31 [-11.95, -4.68] | 26.70 [23.01, 30.40] |
| 1 | 658 | 10984 | 5.53 [5.01-6.08] | 587 | 10932 | 4.78 [4.29-5.31] | 845 | 16980 | 4.31 [3.93-4.72] | 969 | 16593 | 5.35 [4.93-5.79] | -13.56 [-22.78, -4.34] | -9.83 [-19.04, -0.63] | 24.13 [14.85, 33.41] |
| 2 | 233 | 3815 | 5.67 [4.81-6.64] | 212 | 3891 | 4.88 [4.07-5.79] | 296 | 5994 | 4.26 [3.64-4.96] | 372 | 6145 | 5.61 [4.92-6.36] | -13.93 [-29.28, 1.41] | -12.50 [-27.87, 2.87] | 31.22 [15.49, 46.95] |
| >2 | 29 | 415 | 6.73 [4.23-10.19] | 32 | 383 | 8.38 [5.52-12.24] | 27 | 643 | 3.37 [1.81-5.59] | 38 | 639 | 5.48 [3.57-8.02] | 24.67 [-20.95, 70.28] | -59.90 [-91.41, -28.40] | 62.61 [3.56, 121.66] |
| **Ethnicity** (granular)** |  |  |  |  |  |  |  |  |  |  |  |  |  |  |  |
| White British | 4415 | 86790 | 4.44 [4.27-4.62] | 4017 | 92292 | 3.56 [3.40-3.72] | 5665 | 139305 | 3.21 [3.09-3.34] | 6856 | 141124 | 4.17 [4.03-4.30] | -20.27 [-23.87, -16.67] | -9.27 [-13.20, -5.34] | 29.60 [25.55, 33.64] |
| White Irish | 78 | 968 | 8.02 [6.15-10.29] | 64 | 990 | 6.10 [4.45-8.16] | 85 | 1538 | 4.97 [3.72-6.49] | 108 | 1519 | 6.88 [5.45-8.57] | -23.94 [-47.76, -0.12] | -18.36 [-44.43, 7.70] | 38.23 [9.46, 67.00] |
| White gypsy or Irish traveller | 1 | 32 | 2.08 [0.00-17.28] | 2 | 21 | 9.79 [1.51-33.15] | 3 | 33 | 9.27 [2.10-26.71] | 1 | 35 | 1.76 [0.00-15.82] | 370.67 [-185.10, 926.44] | -5.21 [-140.25, 129.83] | -81.01 [-181.77, 19.74] |
| Any other white background | 333 | 4947 | 6.42 [5.62-7.31] | 301 | 4700 | 6.03 [5.23-6.92] | 423 | 7351 | 5.25 [4.63-5.92] | 485 | 7849 | 5.76 [5.14-6.43] | -6.23 [-19.00, 6.54] | -12.94 [-25.04, -0.83] | 9.90 [-1.90, 21.71] |
| Mixed white / black Carribean | 27 | 321 | 8.45 [5.36-12.72] | 17 | 313 | 4.86 [2.43-8.58] | 23 | 436 | 4.67 [2.58-7.70] | 35 | 427 | 8.19 [5.48-11.79] | -42.49 [-81.18, -3.79] | -3.91 [-59.88, 52.06] | 75.37 [15.85, 134.90] |
| mixed white / black African | 12 | 107 | 11.83 [6.18-20.71] | 8 | 110 | 7.08 [2.81-14.82] | 8 | 163 | 4.23 [1.33-9.62] | 12 | 179 | 6.39 [2.98-11.99] | -40.15 [-94.17, 13.86] | -40.11 [-109.04, 28.81] | 51.06 [-45.86, 147.99] |
| Mixed white and Asian | 26 | 396 | 6.22 [3.76-9.69] | 24 | 422 | 5.17 [2.95-8.34] | 32 | 608 | 4.65 [2.83-7.15] | 51 | 682 | 7.32 [5.22-10.00] | -17.04 [-60.93, 26.85] | -10.06 [-55.51, 35.40] | 57.63 [10.32, 104.95] |
| Any other mixed / multiple ethnic background | 41 | 523 | 7.76 [5.34-10.92] | 27 | 463 | 5.34 [3.18-8.38] | 45 | 658 | 6.55 [4.52-9.19] | 42 | 671 | 5.85 [3.93-8.38] | -31.19 [-64.82, 2.45] | 22.66 [-22.10, 67.42] | -10.53 [-44.27, 23.21] |
| Indian | 177 | 1847 | 9.86 [8.34-11.58] | 177 | 2092 | 8.51 [7.16-10.04] | 212 | 2920 | 7.06 [5.99-8.26] | 260 | 2896 | 9.13 [7.94-10.45] | -13.69 [-28.80, 1.42] | -17.04 [-31.84, -2.23] | 29.32 [12.89, 45.75] |
| Pakistani | 54 | 503 | 11.25 [8.34-14.87] | 52 | 509 | 10.62 [7.80-14.16] | 73 | 720 | 10.53 [8.12-13.45] | 89 | 686 | 13.94 [11.16-17.22] | -5.51 [-33.33, 22.31] | -0.94 [-27.78, 25.89] | 32.38 [5.98, 58.78] |
| Bangladeshi | 24 | 189 | 13.61 [8.78-20.24] | 27 | 224 | 12.84 [8.45-18.76] | 26 | 289 | 9.15 [5.80-13.80] | 41 | 294 | 15.12 [10.90-20.45] | -5.73 [-44.67, 33.21] | -28.58 [-63.40, 6.23] | 65.14 [18.36, 111.91] |
| Chinese | 29 | 410 | 6.84 [4.30-10.33] | 18 | 406 | 3.65 [1.71-6.62] | 30 | 614 | 4.20 [2.46-6.61] | 38 | 608 | 5.84 [3.84-8.51] | -46.49 [-85.23, -7.75] | 15.07 [-44.66, 74.79] | 39.05 [-11.67, 89.76] |
| Other Asian | 85 | 709 | 12.76 [10.12-15.89] | 66 | 699 | 9.69 [7.34-12.57] | 98 | 975 | 10.42 [8.33-12.89] | 99 | 941 | 10.99 [8.82-13.55] | -24.06 [-45.06, -3.06] | 7.53 [-17.13, 32.20] | 5.47 [-16.22, 27.16] |
| African | 90 | 519 | 19.21 [15.57-23.42] | 69 | 535 | 13.85 [10.75-17.60] | 108 | 749 | 15.69 [12.87-18.94] | 84 | 668 | 13.46 [10.69-16.75] | -27.80 [-46.49, -9.11] | 13.21 [-9.53, 35.96] | -14.21 [-33.01, 4.59] |
| Caribbean | 38 | 320 | 12.62 [8.89-17.45] | 27 | 322 | 8.42 [5.33-12.68] | 29 | 444 | 6.18 [3.84-9.43] | 29 | 414 | 6.75 [4.24-10.22] | -33.20 [-63.87, -2.54] | -26.60 [-64.13, 10.93] | 9.22 [-35.92, 54.37] |
| Other black / African / Caribbean background | 7 | 61 | 12.14 [5.15-24.63] | 6 | 79 | 7.46 [2.56-17.10] | 17 | 111 | 16.77 [10.11-26.22] | 6 | 98 | 5.69 [1.73-13.64] | -38.55 [-106.26, 29.16] | 124.80 [25.87, 223.73] | -66.07 [-106.74, -25.40] |
| Arab | 15 | 168 | 9.07 [4.92-15.43] | 10 | 169 | 5.44 [2.22-11.02] | 20 | 248 | 8.03 [4.68-12.92] | 24 | 232 | 10.78 [6.81-16.30] | -39.91 [-91.07, 11.25] | 47.24 [-27.76, 122.24] | 34.25 [-19.30, 87.80] |
| Any other ethnic group | 64 | 594 | 11.29 [8.59-14.61] | 60 | 770 | 7.70 [5.66-10.24] | 78 | 1145 | 6.52 [4.93-8.46] | 108 | 1083 | 10.33 [8.35-12.65] | -31.80 [-54.83, -8.77] | -15.32 [-41.04, 10.39] | 58.44 [29.14, 87.73] |
| Prefer not to say | 28 | 479 | 5.36 [3.22-8.34] | 23 | 713 | 2.20 [0.91-4.09] | 61 | 1035 | 5.41 [3.87-7.35] | 63 | 1130 | 5.03 [3.59-6.83] | -58.96 [-97.20, -20.71] | 145.91 [72.73, 219.09] | -6.84 [-36.97, 23.29] |
| **Employment (broad)** |  |  |  |  |  |  |  |  |  |  |  |  |  |  |  |
| Healthcare | 452 | 4553 | 10.27 [9.27-11.36] | 451 | 4623 | 10.07 [9.08-11.14] | 691 | 7108 | 10.03 [9.22-10.88] | 722 | 7996 | 9.19 [8.46-9.97] | -2.04 [-11.88, 7.79] | -0.50 [-9.53, 8.54] | -8.28 [-15.95, -0.60] |
| Care home | 127 | 907 | 15.18 [12.65-18.09] | 95 | 951 | 10.35 [8.24-12.84] | 131 | 1236 | 11.08 [9.16-13.30] | 115 | 1086 | 11.07 [9.03-13.45] | -31.82 [-47.96, -15.68] | 7.05 [-13.53, 27.63] | -0.09 [-18.86, 18.68] |
| Other essential worker | 1209 | 19927 | 5.62 [5.23-6.03] | 1019 | 19615 | 4.57 [4.21-4.96] | 1463 | 29572 | 4.27 [3.98-4.58] | 1799 | 31787 | 5.13 [4.83-5.44] | -18.68 [-25.44, -11.92] | -6.56 [-13.79, 0.66] | 19.91 [13.11, 26.70] |
| Other worker | 2189 | 37855 | 5.28 [5.00-5.57] | 1982 | 40782 | 4.17 [3.92-4.43] | 2704 | 60731 | 3.68 [3.48-3.88] | 3197 | 59632 | 4.77 [4.56-4.99] | -21.02 [-25.95, -16.10] | -11.99 [-17.27, -6.71] | 29.89 [24.46, 35.33] |
| Not in employment | 1516 | 35737 | 3.42 [3.18-3.68] | 1412 | 39030 | 2.67 [2.45-2.90] | 1988 | 59369 | 2.35 [2.18-2.53] | 2500 | 59546 | 3.37 [3.18-3.57] | -21.93 [-28.65, -15.20] | -11.99 [-19.48, -4.49] | 43.40 [35.74, 51.06] |
| **Age group** (broad)** |  |  |  |  |  |  |  |  |  |  |  |  |  |  |  |
| 18-24 | 463 | 6499 | 6.90 [6.17-7.68] | 411 | 6493 | 5.94 [5.26-6.68] | 574 | 8763 | 6.21 [5.60-6.85] | 851 | 9539 | 9.06 [8.39-9.77] | -13.77 [-24.06, -3.48] | 4.38 [-6.57, 15.32] | 46.05 [35.75, 56.36] |
| 25-44 | 1894 | 30418 | 5.82 [5.49-6.15] | 1612 | 30703 | 4.64 [4.35-4.95] | 2238 | 46899 | 4.06 [3.83-4.30] | 2652 | 47933 | 4.98 [4.74-5.23] | -20.27 [-25.60, -14.95] | -12.50 [-18.10, -6.90] | 22.66 [17.00, 28.33] |
| 45-64 | 2386 | 41038 | 5.32 [5.05-5.60] | 2174 | 43327 | 4.36 [4.12-4.61] | 3096 | 65273 | 4.03 [3.83-4.23] | 3579 | 66095 | 4.84 [4.63-5.05] | -17.86 [-22.56, -13.16] | -7.80 [-12.84, -2.75] | 20.10 [15.14, 25.06] |
| 65+ | 801 | 21953 | 2.71 [2.42-3.02] | 798 | 25306 | 2.11 [1.86-2.38] | 1129 | 38432 | 1.85 [1.65-2.06] | 1349 | 37970 | 2.59 [2.38-2.82] | -22.14 [-32.10, -12.18] | -11.85 [-22.75, -0.95] | 39.46 [28.11, 50.81] |

### Figure S1: REACT-2 Round 4: SARS CoV-2 antibody prevalence by Lower tier local authority area (27 Oct – 10 Nov 2020, n=161537)

### Data table below

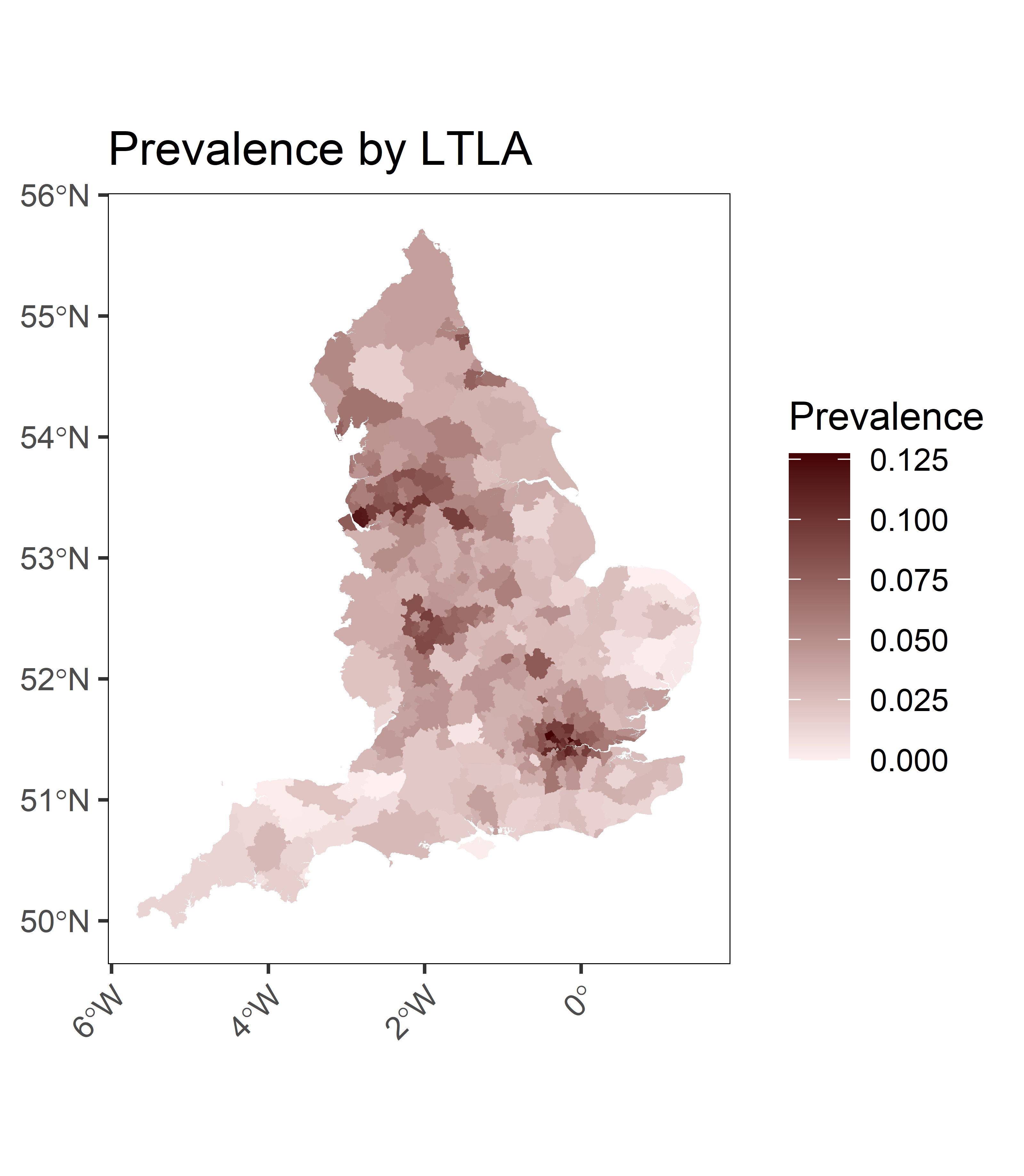

### Table S5: REACT-2 Round 4: Prevalence by lower tier local authority

Prevalence by local authority district (n = 161537).

| **District** | **Positive** | **Total** | **Prevalence %** | **95% CI Lower** | **95% CI Upper** |
| --- | --- | --- | --- | --- | --- |
| Adur | 17 | 502 | 2.39 | 0.87 | 4.77 |
| Allerdale | 30 | 520 | 5.26 | 3.22 | 8.09 |
| Amber Valley | 24 | 508 | 4.01 | 2.16 | 6.67 |
| Arun | 15 | 529 | 1.73 | 0.39 | 3.89 |
| Ashfield | 24 | 549 | 3.58 | 1.87 | 6.05 |
| Ashford | 19 | 513 | 2.78 | 1.19 | 5.2 |
| Aylesbury Vale | 23 | 563 | 3.24 | 1.61 | 5.61 |
| Babergh | 10 | 527 | 0.6 | 0 | 2.48 |
| Barking and Dagenham | 31 | 381 | 8.12 | 5.29 | 11.9 |
| Barnet | 45 | 496 | 9.24 | 6.57 | 12.7 |
| Barnsley | 31 | 525 | 5.43 | 3.36 | 8.26 |
| Barrow-in-Furness | 38 | 501 | 7.45 | 5.03 | 10.6 |
| Basildon | 31 | 498 | 5.81 | 3.64 | 8.8 |
| Basingstoke and Deane | 17 | 541 | 2.1 | 0.69 | 4.31 |
| Bassetlaw | 30 | 508 | 5.43 | 3.33 | 8.32 |
| Bath and North East Somerset | 21 | 549 | 2.92 | 1.34 | 5.27 |
| Bedford | 38 | 481 | 7.83 | 5.31 | 11.2 |
| Bexley | 41 | 440 | 9.54 | 6.68 | 13.2 |
| Birmingham | 42 | 492 | 8.6 | 6 | 12 |
| Blaby | 30 | 498 | 5.57 | 3.44 | 8.52 |
| Blackburn with Darwen | 35 | 464 | 7.4 | 4.91 | 10.7 |
| Blackpool | 31 | 469 | 6.28 | 3.97 | 9.43 |
| Bolsover | 19 | 492 | 2.97 | 1.31 | 5.48 |
| Bolton | 38 | 487 | 7.71 | 5.23 | 11 |
| Boston | 18 | 486 | 2.78 | 1.15 | 5.27 |
| Bournemouth, Christchurch and Poole | 19 | 527 | 2.66 | 1.11 | 5.01 |
| Bracknell Forest | 22 | 524 | 3.37 | 1.67 | 5.87 |
| Bradford | 36 | 517 | 6.7 | 4.42 | 9.75 |
| Braintree | 21 | 523 | 3.15 | 1.5 | 5.61 |
| Breckland | 13 | 484 | 1.55 | 0.21 | 3.78 |
| Brent | 45 | 406 | 11.7 | 8.42 | 15.8 |
| Brentwood | 33 | 478 | 6.63 | 4.29 | 9.8 |
| Brighton and Hove | 14 | 549 | 1.39 | 0.15 | 3.42 |
| Bristol, City of | 23 | 501 | 3.84 | 2.02 | 6.5 |
| Broadland | 11 | 545 | 0.74 | 0 | 2.62 |
| Bromley | 38 | 518 | 7.15 | 4.81 | 10.2 |
| Bromsgrove | 51 | 583 | 8.85 | 6.4 | 11.9 |
| Broxbourne | 33 | 463 | 6.9 | 4.48 | 10.2 |
| Broxtowe | 23 | 572 | 3.16 | 1.56 | 5.5 |
| Burnley | 38 | 451 | 8.46 | 5.78 | 12 |
| Bury | 31 | 517 | 5.54 | 3.44 | 8.42 |
| Calderdale | 42 | 521 | 8.03 | 5.57 | 11.2 |
| Cambridge | 22 | 576 | 2.91 | 1.37 | 5.2 |
| Camden | 53 | 526 | 10.4 | 7.69 | 13.9 |
| Cannock Chase | 25 | 499 | 4.35 | 2.43 | 7.1 |
| Canterbury | 19 | 510 | 2.8 | 1.2 | 5.24 |
| Carlisle | 24 | 515 | 3.93 | 2.11 | 6.55 |
| Castle Point | 30 | 454 | 6.27 | 3.94 | 9.49 |
| Central Bedfordshire | 22 | 516 | 3.45 | 1.73 | 5.99 |
| Charnwood | 21 | 532 | 3.07 | 1.44 | 5.49 |
| Chelmsford | 19 | 548 | 2.49 | 1 | 4.76 |
| Cheltenham | 30 | 578 | 4.57 | 2.72 | 7.12 |
| Cherwell | 29 | 552 | 4.64 | 2.75 | 7.28 |
| Cheshire East | 30 | 534 | 5.08 | 3.09 | 7.84 |
| Cheshire West and Chester | 21 | 525 | 3.13 | 1.48 | 5.59 |
| Chesterfield | 20 | 523 | 2.92 | 1.31 | 5.34 |
| Chichester | 15 | 559 | 1.55 | 0.28 | 3.59 |
| Chiltern | 33 | 565 | 5.35 | 3.36 | 8.06 |
| Chorley | 40 | 517 | 7.63 | 5.22 | 10.8 |
| Colchester | 26 | 560 | 3.91 | 2.15 | 6.4 |
| Copeland | 23 | 488 | 3.99 | 2.12 | 6.71 |
| Corby | 20 | 455 | 3.61 | 1.76 | 6.38 |
| Cornwall | 13 | 512 | 1.37 | 0.11 | 3.49 |
| Cotswold | 29 | 546 | 4.71 | 2.8 | 7.38 |
| County Durham | 22 | 522 | 3.39 | 1.69 | 5.9 |
| Coventry | 26 | 498 | 4.6 | 2.64 | 7.4 |
| Craven | 30 | 573 | 4.62 | 2.76 | 7.2 |
| Crawley | 15 | 441 | 2.41 | 0.81 | 4.98 |
| Croydon | 30 | 481 | 5.83 | 3.62 | 8.87 |
| Dacorum | 24 | 558 | 3.5 | 1.82 | 5.93 |
| Darlington | 22 | 471 | 3.94 | 2.05 | 6.71 |
| Dartford | 37 | 493 | 7.36 | 4.93 | 10.6 |
| Daventry | 22 | 520 | 3.41 | 1.7 | 5.93 |
| Derby | 32 | 557 | 5.24 | 3.25 | 7.95 |
| Derbyshire Dales | 23 | 578 | 3.11 | 1.53 | 5.42 |
| Doncaster | 29 | 499 | 5.32 | 3.22 | 8.22 |
| Dorset | 22 | 598 | 2.75 | 1.26 | 4.95 |
| Dover | 16 | 500 | 2.17 | 0.7 | 4.5 |
| Dudley | 40 | 502 | 7.91 | 5.43 | 11.2 |
| Ealing | 36 | 493 | 7.11 | 4.72 | 10.3 |
| East Cambridgeshire | 19 | 555 | 2.44 | 0.97 | 4.68 |
| East Devon | 12 | 536 | 1.01 | 0 | 2.98 |
| East Hampshire | 17 | 521 | 2.24 | 0.78 | 4.53 |
| East Hertfordshire | 31 | 522 | 5.47 | 3.39 | 8.32 |
| East Lindsey | 19 | 522 | 2.7 | 1.14 | 5.08 |
| East Northamptonshire | 18 | 512 | 2.55 | 1.01 | 4.93 |
| East Riding of Yorkshire | 21 | 548 | 2.93 | 1.35 | 5.29 |
| East Staffordshire | 23 | 472 | 4.18 | 2.25 | 6.99 |
| East Suffolk | 9 | 504 | 0.46 | 0 | 2.36 |
| Eastbourne | 20 | 507 | 3.07 | 1.41 | 5.56 |
| Eastleigh | 23 | 573 | 3.15 | 1.55 | 5.48 |
| Eden | 14 | 505 | 1.65 | 0.31 | 3.85 |
| Elmbridge | 34 | 535 | 5.97 | 3.84 | 8.85 |
| Enfield | 49 | 491 | 10.3 | 7.51 | 13.9 |
| Epping Forest | 29 | 453 | 6.03 | 3.73 | 9.21 |
| Epsom and Ewell | 38 | 533 | 6.9 | 4.62 | 9.92 |
| Erewash | 25 | 549 | 3.8 | 2.05 | 6.31 |
| Exeter | 15 | 571 | 1.48 | 0.24 | 3.48 |
| Fareham | 15 | 534 | 1.7 | 0.37 | 3.83 |
| Fenland | 16 | 507 | 2.12 | 0.66 | 4.42 |
| Folkestone and Hythe | 13 | 531 | 1.26 | 0.04 | 3.31 |
| Forest of Dean | 12 | 488 | 1.28 | 0.01 | 3.43 |
| Fylde | 28 | 536 | 4.61 | 2.7 | 7.28 |
| Gateshead | 28 | 470 | 5.49 | 3.32 | 8.52 |
| Gedling | 23 | 538 | 3.46 | 1.77 | 5.94 |
| Gloucester | 27 | 536 | 4.38 | 2.51 | 7.02 |
| Gosport | 19 | 576 | 2.29 | 0.87 | 4.45 |
| Gravesham | 28 | 484 | 5.28 | 3.17 | 8.23 |
| Great Yarmouth | 10 | 437 | 1.07 | 0 | 3.33 |
| Greenwich | 32 | 482 | 6.31 | 4.03 | 9.42 |
| Guildford | 23 | 546 | 3.39 | 1.71 | 5.83 |
| Hackney | 59 | 498 | 12.6 | 9.51 | 16.4 |
| Halton | 23 | 384 | 5.53 | 3.16 | 8.95 |
| Hambleton | 22 | 555 | 3.09 | 1.48 | 5.46 |
| Hammersmith and Fulham | 54 | 515 | 10.9 | 8.1 | 14.5 |
| Harborough | 19 | 528 | 2.65 | 1.1 | 5 |
| Haringey | 43 | 469 | 9.36 | 6.6 | 12.9 |
| Harlow | 26 | 474 | 4.92 | 2.86 | 7.85 |
| Harrogate | 33 | 541 | 5.66 | 3.59 | 8.49 |
| Harrow | 63 | 526 | 12.7 | 9.73 | 16.4 |
| Hart | 16 | 536 | 1.91 | 0.54 | 4.09 |
| Hartlepool | 26 | 473 | 4.94 | 2.87 | 7.87 |
| Hastings | 16 | 505 | 2.13 | 0.67 | 4.44 |
| Havant | 14 | 508 | 1.63 | 0.3 | 3.82 |
| Havering | 36 | 463 | 7.68 | 5.14 | 11.1 |
| Herefordshire, County of | 17 | 519 | 2.26 | 0.79 | 4.56 |
| Hertsmere | 47 | 514 | 9.33 | 6.68 | 12.7 |
| High Peak | 25 | 539 | 3.9 | 2.12 | 6.45 |
| Hillingdon | 39 | 467 | 8.37 | 5.75 | 11.8 |
| Hinckley and Bosworth | 38 | 538 | 6.82 | 4.57 | 9.81 |
| Horsham | 16 | 540 | 1.88 | 0.52 | 4.05 |
| Hounslow | 47 | 461 | 10.6 | 7.66 | 14.3 |
| Huntingdonshire | 19 | 533 | 2.61 | 1.08 | 4.94 |
| Hyndburn | 36 | 435 | 8.28 | 5.59 | 11.9 |
| Ipswich | 19 | 510 | 2.8 | 1.2 | 5.24 |
| Isle of Wight | 8 | 521 | 0.16 | 0 | 1.93 |
| Islington | 46 | 548 | 8.43 | 5.97 | 11.6 |
| Kensington and Chelsea | 47 | 531 | 8.98 | 6.41 | 12.3 |
| Kettering | 14 | 497 | 1.71 | 0.34 | 3.94 |
| King's Lynn and West Norfolk | 18 | 518 | 2.5 | 0.97 | 4.85 |
| Kingston upon Hull, City of | 20 | 483 | 3.3 | 1.56 | 5.91 |
| Kingston upon Thames | 42 | 548 | 7.55 | 5.2 | 10.6 |
| Kirklees | 41 | 531 | 7.62 | 5.23 | 10.7 |
| Knowsley | 50 | 456 | 11.5 | 8.45 | 15.4 |
| Lambeth | 57 | 536 | 11.1 | 8.31 | 14.6 |
| Lancaster | 27 | 509 | 4.7 | 2.74 | 7.48 |
| Leeds | 27 | 529 | 4.46 | 2.57 | 7.14 |
| Leicester | 27 | 405 | 6.35 | 3.88 | 9.79 |
| Lewes | 20 | 575 | 2.5 | 1.04 | 4.71 |
| Lewisham | 49 | 507 | 9.96 | 7.21 | 13.4 |
| Lichfield | 29 | 555 | 4.61 | 2.73 | 7.23 |
| Lincoln | 22 | 536 | 3.26 | 1.6 | 5.71 |
| Liverpool | 48 | 429 | 11.8 | 8.61 | 15.8 |
| Luton | 39 | 479 | 8.12 | 5.56 | 11.5 |
| Maidstone | 13 | 517 | 1.34 | 0.09 | 3.44 |
| Maldon | 19 | 494 | 2.95 | 1.3 | 5.46 |
| Malvern Hills | 26 | 560 | 3.91 | 2.15 | 6.4 |
| Manchester | 50 | 524 | 9.81 | 7.12 | 13.2 |
| Mansfield | 23 | 569 | 3.18 | 1.58 | 5.53 |
| Medway | 17 | 484 | 2.55 | 0.97 | 5 |
| Melton | 34 | 547 | 5.8 | 3.71 | 8.63 |
| Mendip | 7 | 520 | 0 | 0 | 1.63 |
| Merton | 47 | 508 | 9.46 | 6.78 | 12.9 |
| Mid Devon | 9 | 561 | 0.25 | 0 | 1.95 |
| Mid Suffolk | 8 | 523 | 0.16 | 0 | 1.91 |
| Mid Sussex | 17 | 498 | 2.43 | 0.89 | 4.82 |
| Middlesbrough | 36 | 503 | 6.94 | 4.6 | 10.1 |
| Milton Keynes | 25 | 506 | 4.27 | 2.37 | 6.98 |
| Mole Valley | 34 | 514 | 6.28 | 4.06 | 9.28 |
| New Forest | 15 | 528 | 1.74 | 0.4 | 3.9 |
| Newark and Sherwood | 22 | 514 | 3.47 | 1.74 | 6.02 |
| Newcastle upon Tyne | 31 | 523 | 5.45 | 3.38 | 8.3 |
| Newcastle-under-Lyme | 23 | 513 | 3.71 | 1.93 | 6.31 |
| Newham | 29 | 274 | 11.1 | 7.31 | 16.1 |
| North Devon | 8 | 503 | 0.23 | 0 | 2.06 |
| North East Derbyshire | 29 | 536 | 4.83 | 2.88 | 7.54 |
| North East Lincolnshire | 18 | 471 | 2.92 | 1.24 | 5.49 |
| North Hertfordshire | 28 | 558 | 4.36 | 2.52 | 6.94 |
| North Kesteven | 18 | 547 | 2.28 | 0.83 | 4.51 |
| North Lincolnshire | 21 | 481 | 3.57 | 1.77 | 6.24 |
| North Norfolk | 7 | 536 | 0 | 0 | 1.53 |
| North Somerset | 20 | 547 | 2.72 | 1.18 | 5.04 |
| North Tyneside | 23 | 506 | 3.79 | 1.99 | 6.42 |
| North Warwickshire | 36 | 484 | 7.27 | 4.84 | 10.5 |
| North West Leicestershire | 23 | 537 | 3.47 | 1.77 | 5.96 |
| Northampton | 34 | 468 | 7.07 | 4.63 | 10.3 |
| Northumberland | 25 | 522 | 4.08 | 2.25 | 6.72 |
| Norwich | 21 | 508 | 3.29 | 1.59 | 5.83 |
| Nottingham | 27 | 487 | 4.99 | 2.94 | 7.89 |
| Nuneaton and Bedworth | 33 | 478 | 6.63 | 4.29 | 9.8 |
| Oadby and Wigston | 25 | 445 | 5.08 | 2.93 | 8.15 |
| Oldham | 43 | 445 | 9.96 | 7.05 | 13.7 |
| Oxford | 25 | 541 | 3.88 | 2.11 | 6.42 |
| Pendle | 27 | 461 | 5.37 | 3.2 | 8.42 |
| Peterborough | 28 | 513 | 4.89 | 2.9 | 7.68 |
| Plymouth | 10 | 488 | 0.78 | 0 | 2.81 |
| Portsmouth | 24 | 491 | 4.2 | 2.3 | 6.95 |
| Preston | 33 | 482 | 6.56 | 4.24 | 9.71 |
| Reading | 31 | 564 | 4.94 | 3.01 | 7.59 |
| Redbridge | 36 | 442 | 8.13 | 5.47 | 11.6 |
| Redcar and Cleveland | 36 | 531 | 6.48 | 4.26 | 9.45 |
| Redditch | 35 | 489 | 6.94 | 4.57 | 10.1 |
| Reigate and Banstead | 22 | 505 | 3.56 | 1.8 | 6.15 |
| Ribble Valley | 24 | 500 | 4.1 | 2.22 | 6.8 |
| Richmond upon Thames | 29 | 553 | 4.63 | 2.74 | 7.26 |
| Richmondshire | 22 | 537 | 3.25 | 1.59 | 5.69 |
| Rochdale | 33 | 477 | 6.65 | 4.3 | 9.83 |
| Rochford | 28 | 506 | 4.98 | 2.96 | 7.81 |
| Rossendale | 31 | 450 | 6.61 | 4.21 | 9.89 |
| Rother | 13 | 516 | 1.35 | 0.09 | 3.45 |
| Rotherham | 35 | 531 | 6.25 | 4.07 | 9.19 |
| Rugby | 19 | 517 | 2.74 | 1.16 | 5.14 |
| Runnymede | 13 | 533 | 1.25 | 0.04 | 3.29 |
| Rushcliffe | 33 | 588 | 5.07 | 3.16 | 7.68 |
| Rushmoor | 12 | 504 | 1.18 | 0 | 3.27 |
| Rutland | 17 | 504 | 2.38 | 0.86 | 4.74 |
| Ryedale | 22 | 528 | 3.33 | 1.65 | 5.82 |
| Salford | 45 | 453 | 10.3 | 7.36 | 14 |
| Sandwell | 29 | 434 | 6.36 | 3.97 | 9.68 |
| Scarborough | 20 | 552 | 2.68 | 1.15 | 4.98 |
| Sedgemoor | 8 | 483 | 0.31 | 0 | 2.21 |
| Sefton | 35 | 497 | 6.8 | 4.47 | 9.92 |
| Selby | 18 | 543 | 2.31 | 0.85 | 4.55 |
| Sevenoaks | 17 | 538 | 2.12 | 0.7 | 4.34 |
| Sheffield | 50 | 547 | 9.33 | 6.75 | 12.6 |
| Shropshire | 23 | 548 | 3.37 | 1.7 | 5.81 |
| Slough | 32 | 463 | 6.64 | 4.26 | 9.87 |
| Solihull | 40 | 489 | 8.17 | 5.62 | 11.5 |
| Somerset West and Taunton | 18 | 558 | 2.2 | 0.78 | 4.39 |
| South Bucks | 35 | 516 | 6.49 | 4.24 | 9.5 |
| South Cambridgeshire | 18 | 535 | 2.37 | 0.89 | 4.64 |
| South Derbyshire | 25 | 525 | 4.05 | 2.22 | 6.67 |
| South Gloucestershire | 30 | 597 | 4.37 | 2.58 | 6.85 |
| South Hams | 15 | 533 | 1.7 | 0.38 | 3.84 |
| South Holland | 13 | 490 | 1.51 | 0.19 | 3.72 |
| South Kesteven | 19 | 511 | 2.79 | 1.2 | 5.22 |
| South Lakeland | 36 | 536 | 6.41 | 4.21 | 9.35 |
| South Norfolk | 18 | 540 | 2.33 | 0.87 | 4.59 |
| South Northamptonshire | 29 | 550 | 4.67 | 2.77 | 7.31 |
| South Oxfordshire | 22 | 547 | 3.16 | 1.53 | 5.56 |
| South Ribble | 25 | 537 | 3.92 | 2.14 | 6.48 |
| South Somerset | 12 | 554 | 0.92 | 0 | 2.83 |
| South Staffordshire | 44 | 526 | 8.39 | 5.89 | 11.6 |
| South Tyneside | 30 | 491 | 5.67 | 3.51 | 8.66 |
| Southampton | 19 | 497 | 2.92 | 1.28 | 5.41 |
| Southend-on-Sea | 32 | 482 | 6.31 | 4.03 | 9.42 |
| Southwark | 51 | 491 | 10.8 | 7.94 | 14.5 |
| Spelthorne | 29 | 498 | 5.33 | 3.23 | 8.24 |
| St Albans | 30 | 557 | 4.8 | 2.89 | 7.45 |
| St. Helens | 47 | 507 | 9.48 | 6.8 | 12.9 |
| Stafford | 21 | 536 | 3.03 | 1.42 | 5.44 |
| Staffordshire Moorlands | 23 | 508 | 3.77 | 1.97 | 6.39 |
| Stevenage | 18 | 499 | 2.66 | 1.08 | 5.1 |
| Stockport | 38 | 543 | 6.74 | 4.51 | 9.71 |
| Stockton-on-Tees | 40 | 528 | 7.44 | 5.08 | 10.5 |
| Stoke-on-Trent | 25 | 465 | 4.79 | 2.73 | 7.73 |
| Stratford-on-Avon | 17 | 547 | 2.06 | 0.66 | 4.24 |
| Stroud | 25 | 539 | 3.9 | 2.12 | 6.45 |
| Sunderland | 40 | 484 | 8.27 | 5.7 | 11.6 |
| Surrey Heath | 29 | 559 | 4.56 | 2.69 | 7.17 |
| Sutton | 38 | 517 | 7.17 | 4.82 | 10.3 |
| Swale | 18 | 510 | 2.57 | 1.02 | 4.95 |
| Swindon | 23 | 533 | 3.51 | 1.8 | 6.01 |
| Tameside | 29 | 456 | 5.98 | 3.69 | 9.14 |
| Tamworth | 32 | 485 | 6.26 | 3.99 | 9.36 |
| Tandridge | 29 | 560 | 4.55 | 2.69 | 7.15 |
| Teignbridge | 13 | 508 | 1.4 | 0.12 | 3.53 |
| Telford and Wrekin | 20 | 487 | 3.26 | 1.53 | 5.85 |
| Tendring | 26 | 535 | 4.17 | 2.33 | 6.78 |
| Test Valley | 18 | 530 | 2.41 | 0.91 | 4.7 |
| Tewkesbury | 28 | 581 | 4.12 | 2.36 | 6.6 |
| Thanet | 17 | 478 | 2.6 | 1 | 5.09 |
| Three Rivers | 42 | 524 | 7.97 | 5.52 | 11.2 |
| Thurrock | 31 | 481 | 6.08 | 3.83 | 9.16 |
| Tonbridge and Malling | 21 | 501 | 3.36 | 1.64 | 5.93 |
| Torbay | 9 | 511 | 0.44 | 0 | 2.3 |
| Torridge | 12 | 506 | 1.17 | 0 | 3.25 |
| Tower Hamlets | 42 | 497 | 8.49 | 5.92 | 11.8 |
| Trafford | 35 | 509 | 6.6 | 4.32 | 9.65 |
| Tunbridge Wells | 21 | 515 | 3.23 | 1.54 | 5.73 |
| Uttlesford | 22 | 524 | 3.37 | 1.67 | 5.87 |
| Vale of White Horse | 10 | 539 | 0.55 | 0 | 2.39 |
| Wakefield | 26 | 515 | 4.4 | 2.49 | 7.1 |
| Walsall | 44 | 476 | 9.45 | 6.7 | 13 |
| Waltham Forest | 34 | 480 | 6.85 | 4.47 | 10 |
| Wandsworth | 49 | 521 | 9.64 | 6.97 | 13 |
| Warrington | 27 | 527 | 4.49 | 2.58 | 7.17 |
| Warwick | 28 | 539 | 4.57 | 2.67 | 7.24 |
| Watford | 33 | 434 | 7.47 | 4.9 | 10.9 |
| Waverley | 17 | 553 | 2.02 | 0.64 | 4.18 |
| Wealden | 14 | 525 | 1.53 | 0.23 | 3.65 |
| Wellingborough | 22 | 471 | 3.94 | 2.05 | 6.71 |
| Welwyn Hatfield | 26 | 521 | 4.33 | 2.44 | 7 |
| West Berkshire | 18 | 559 | 2.19 | 0.78 | 4.38 |
| West Devon | 20 | 538 | 2.79 | 1.23 | 5.15 |
| West Lancashire | 34 | 546 | 5.82 | 3.72 | 8.65 |
| West Lindsey | 13 | 516 | 1.35 | 0.09 | 3.45 |
| West Oxfordshire | 22 | 526 | 3.35 | 1.66 | 5.84 |
| West Suffolk | 11 | 517 | 0.88 | 0 | 2.85 |
| Westminster | 47 | 478 | 10.2 | 7.32 | 13.8 |
| Wigan | 37 | 451 | 8.2 | 5.55 | 11.7 |
| Wiltshire | 17 | 565 | 1.94 | 0.59 | 4.05 |
| Winchester | 25 | 525 | 4.05 | 2.22 | 6.67 |
| Windsor and Maidenhead | 29 | 522 | 5.01 | 3.01 | 7.79 |
| Wirral | 38 | 487 | 7.71 | 5.23 | 11 |
| Woking | 28 | 528 | 4.7 | 2.76 | 7.42 |
| Wokingham | 21 | 573 | 2.73 | 1.22 | 4.99 |
| Wolverhampton | 23 | 433 | 4.71 | 2.61 | 7.76 |
| Worcester | 24 | 523 | 3.84 | 2.05 | 6.43 |
| Worthing | 19 | 487 | 3.01 | 1.34 | 5.56 |
| Wychavon | 34 | 542 | 5.87 | 3.76 | 8.72 |
| Wycombe | 25 | 554 | 3.75 | 2.02 | 6.24 |
| Wyre | 32 | 512 | 5.84 | 3.69 | 8.78 |
| Wyre Forest | 27 | 562 | 4.1 | 2.32 | 6.63 |
| York | 24 | 551 | 3.56 | 1.86 | 6.02 |
